# Quantitative magnetisation transfer imaging in relapsing-remitting multiple sclerosis: a systematic review and meta-analysis

**DOI:** 10.1101/2021.07.14.21260512

**Authors:** Elizabeth N. York, Michael J. Thrippleton, Rozanna Meijboom, David P.J. Hunt, Adam D. Waldman

## Abstract

Myelin-sensitive MRI such as magnetisation transfer imaging has been widely used in the clinical context of multiple sclerosis. The influence of methodology and differences in disease subtype on imaging findings is, however, not well established. Here, we aim to review systematically the use of quantitative magnetisation transfer imaging in the brain in relapsing-remitting multiple sclerosis. We examine how methodological differences, disease effects and their interaction influence magnetisation transfer imaging measures.

Articles published before 06/01/2021 were retrieved from online databases (PubMed, EMBASE and Web of Science) with search terms including ‘magnetisation transfer’ and ‘brain’ for systematic review. Only studies which used human *in vivo* quantitative magnetisation transfer imaging in adults with relapsing-remitting multiple sclerosis (with or without healthy controls) were included.

Data including sample size, magnetic field strength, MRI acquisition protocol parameters, treatments and clinical findings were extracted and qualitatively synthesised. Where possible, effect sizes were calculated for meta-analyses to determine magnetisation transfer (1) differences between patients and healthy controls; (2) longitudinal change; and, (3) relationships with clinical disability in relapsing-remitting multiple sclerosis.

Eighty-six studies met the inclusion/exclusion criteria. MRI acquisition parameters varied widely, and were also underreported. The majority of studies examined MTR (magnetisation transfer ratio) in white matter, but magnetisation transfer metrics, brain regions and results were heterogeneous. Analysis demonstrated a risk of bias due to selective reporting and small sample sizes.

A random-effects meta-analysis revealed MTR was 1.1 percent units [95% CI -1.47pu to -0.73pu] lower in relapsing-remitting multiple sclerosis than healthy controls (z-value: -6.04, p<0.001, n=23). Linear mixed-model analysis did not show a significant longitudinal change in MTR across all brain regions (β=-0.14 [-0.9 to 0.61], t-value=-0.38, p=0.71, n=13) or normal-appearing white matter alone (β=-0.082 [-0.13 to -0.29], t-value=0.78, p=0.44, n=7). There was a significant negative association between MTR and clinical disability, as assessed by the Expanded Disability Status Scale (*r*=-0.30 [95% CI -0.48 to -0.08]; z-value=-2.91, p=0.01, n=8).

Evidence suggests that magnetisation transfer imaging is sensitive to pathological changes in relapsing-remitting multiple sclerosis, although the effect of relapsing-remitting multiple sclerosis on magnetisation transfer metrics in different brain tissue types was small in comparison to the inter-study variability. Recommended improvements include: the use of techniques such as MTsat (magnetisation transfer saturation) or ihMTR (inhomogeneous MTR) which provide more robust and specific microstructural measures within clinically feasible acquisition times; detailed methodological reporting standards; and larger, demographically diverse cohorts for comparison, including healthy controls.

**Abbreviated Summary:** York et al. systematically reviewed 86 studies of magnetisation transfer (MT) brain imaging in relapsing-remitting multiple sclerosis. MT was reduced in patients compared with controls, but results were highly variable, longitudinal change subtle, and associations with clinical disability weak. Use of better harmonised MT acquisition in large cohorts is warranted.

## 1. Introduction

### 1.1. Multiple Sclerosis: a heterogeneous disease

Multiple sclerosis is an immune-mediated disease involving widespread focal injury (lesions) to myelin – the fatty sheath which insulates neuronal axons - and nerve fibres within the CNS, accompanied by neuroinflammation.^1^ This results in irreversible neurodegeneration.

Demyelination and neuronal damage manifest as heterogeneous clinical disability such as weakness, visual disturbances and cognitive impairment. Acute clinical episodes, or relapses, define the relapsing-remitting multiple sclerosis (RRMS) subtype and are often accompanied by new lesions on MRI. Although diverse in pathological appearance, lesions are indicative of inflammation and demyelination. In RRMS, relapses are interspersed with periods of stability or remission, although the clinical course varies and the choice of effective disease-modifying therapies (DMTs) is currently limited.

Reliable, non-invasive *in vivo* biomarkers are necessary to predict and track disease progression in individuals, and objectively assess the effectiveness of both current and emerging treatments.^2^ The relationship between clinical disability and conventional MRI measures of disease burden such as lesion load visible on T2-weighted imaging^3^ and atrophy^4^ is, however, weak. This reflects a need for validated quantitative MRI metrics which are more sensitive and specific to disease-related pathological microstructural change in RRMS.

### 1.2. Magnetisation Transfer Imaging (MTI)

MTI is sensitive to subtle pathological changes in tissue microstructure which cannot typically be quantified with conventional MRI. MT signal is indirectly derived from protons ‘bound’ to macromolecules.

Considering a simple two-pool model for hydrogen nuclei in the brain,^5^ the so-called ‘free’ pool of water protons show relatively unrestricted diffusion and contributes to the bulk source of conventional MRI signal. Hydrogen nuclei in the ‘bound’ pool, however, are closely coupled to macromolecules (including lipids such as myelin) and have hindered rotational and translational motion, resulting in T2 decays too rapid (∼10µs) for the signal to be detectable at typical echo times.

MTI exploits the continuous exchange of magnetisation between pools to obtain signal indirectly from this ‘bound’ pool. Since the frequency spectrum of the ‘bound’ pool is much broader than the ‘free’ water peak, an applied off-resonance radiofrequency pulse may selectively saturate ‘bound’ protons.

Magnetisation exchange between the two pools reduces longitudinal magnetisation of the ‘free’ pool and hence it’s signal intensity. Amongst other factors, the magnitude of this effect depends on the size of the ‘bound’ pool, which hence provides a surrogate marker of myelin integrity. MTI has therefore been used to study white matter diseases, including multiple sclerosis.

### 1.3. Quantifying magnetisation transfer

MTR (magnetisation transfer ratio), calculated as the percentage change in signal with and without a saturation pulse (Video 1), has been widely applied in clinical studies due to relatively brief acquisition and ease of calculation.

MTR is, however, susceptible to field inhomogeneities and T1 relaxation effects, and varies widely depending upon specific acquisition parameters (e.g. TR [repetition time], excitation flip angle, sequence type, saturation pulse offset, power, shape and duration).^6^ Biological interpretation of MTR, as well as inter-site and inter-study comparisons, are therefore challenging, and present a barrier to clinical translation.

MTsat inherently corrects for B1 inhomogeneities and T1 relaxation,^7^ by approximating the signal amplitude and T1 relaxation at low flip angles with an additional T1-weighted image.^7, 8^ MTsat hence addresses some limitations of MTR, within clinically feasible acquisition times and SAR (specific absorption rate) limits, and the resulting parametric maps have visibly better tissue contrast than MTR (Video 1).^7^

ihMTR exploits observed asymmetry of the broadened spectral line of the bound pool, thought to be driven by dipolar coupling effects,^9^ and compares single frequency saturation at positive and negative frequency offsets with simultaneous saturation at two frequencies (+/-).^10, 11^ While not yet fully understood, ihMTR^11^ is thought to be particularly sensitive to highly restricted protons in lipid chains and therefore more specific to the phospholipid bilayer of myelin than other MTI methods.

Fully quantitative MTI (qMT) approaches using multi-compartmental models describe MT effects most rigorously by systematically varying the saturation offset and power. Important derived parameters include the fractional pool size ratio (F, or PSR), the relative macromolecular content (MMC) and the macromolecular proton fraction (*f*) which provide indicators of myelin content. Calculation of either F or *f* requires estimation of the longitudinal relaxation rate, R1, for each pool.^12^ The MT exchange rate from the bound to the free pool (kf) may also help to gauge myelin status. qMT is time-consuming to acquire, requires complex analysis and tends not to provide whole brain coverage. qMT application has therefore mostly been limited to small-scale methodological studies.

### 1.4. Rationale

Previous reviews provide an overview of qMT, MTI^13^ and its specific application in MS.^14–16^ More recently, Weiskopf et al.^17^ have provided a technical review of the concepts, validation and modelling of quantitative MRI, including qMT. The biophysical models used to describe MT effects in tissue, experimental evidence in brain development, aging and pathology have also been reviewed^18^. Lazari and Lipp^19^ and van der Weijden et al.^20^ systematically reviewed myelin-sensitive MRI validation, reproducibility and correlation with histology in humans and animal populations. Campbell et al.^21^ and Mohammadi and Callaghan^22^ have addressed incorporation of MTI-derived g-ratio measures to determine relative myelin-to-axon thickness.

The emergence of methods such as MTsat and ihMTR that provide more specific measures of tissue microstructure than MTR but can be acquired relatively rapidly across the whole brain present an opportunity to reassess the use of clinical MTI.^7, 11, 23^ An evaluation of the body of evidence for MTI as a marker of disease from diverse studies would allow better understanding of the effects of technique and other sources of bias across apparently contradictory results in the literature. Moreover, the differences in clinical course^24^, current therapeutic approaches^25–27^ and CSF diagnostic biomarkers^28^ between multiple sclerosis subtypes justify specific examination in RRMS. We believe therefore that a systematic review of myelin-sensitive MTI in RRMS with meta-analyses is warranted.

### 1.5. Purpose

The aim of the present study is thus to systematically review, in RRMS, (1) MTI techniques used to assess pathological change, and (2) sources of inter-study variability and bias. We then aim to apply meta-analyses to provide consensus on (3) key cross-sectional and longitudinal pathological findings and (4) the relationship between MTI and clinical disability in RRMS.

## 2. Materials and Methods

Approval from an ethics committee was not required for the present review.

### 2.1. Search Strategy & Eligibility Criteria

This review adhered to PRISMA guidelines as far as possible.^29^ The search terms were ‘magnetisation transfer’ or ‘magnetization transfer’ and ‘brain’ (with MeSH terms). The online databases searched were PubMed, Embase and Web of Science.

For inclusion, studies had to be primary human research and had to include people with RRMS. When a study included people with other multiple sclerosis sub-types (e.g. primary progressive) or post-mortem imaging data, it was excluded from the analysis. Articles in any language were accepted, with a publishing cut-off date of 06/01/2021.

Exclusion criteria were: inclusion of subjects with non-MS pathology (e.g. brain tumours, traumatic brain injury); paediatric (i.e. <18 years of age) or paediatric-onset MS; healthy participants only; the full text was not retrievable; only phantom, *in vitro*, preclinical *in vivo* or *ex vivo* data; study published before 1980; an imaging technique other than MTI used; ex- cerebral imaging only; non-quantitative methodology; theoretical or simulation-only papers; a clinical trial protocol, phase I or phase II clinical trial; conference proceedings; a review or opinion article; and, any study clearly irrelevant to the current review. Studies carried out on the same or similar cohorts were not excluded (since this was not typically explicitly clear).

### 2.2. Search Procedure

Search results were imported into EndNote. Duplicate publications were automatically removed using the in-built de-duplicator tool, and remaining duplicates were removed manually. Abstracts were checked by the author (E.Y.) and removed when exclusion criteria were met. Full texts were manually retrieved by the author (E.Y.) with online searches for article DOIs, PMID or title. If this failed, the abstract was excluded. Full-text articles were screened manually by the author (E.Y.) for exclusion criteria and rejected where necessary. The remaining selection were categorised according to the multiple sclerosis subtype. Articles without RRMS cohorts or with mixed subtypes were excluded. Thus, the final selection consisted of studies which recruited only RRMS patients, with or without healthy control subjects.

### 2.3. Data Extraction

Data were extracted in detail including demographics, acquisition parameters, MT measure and brain region, statistical methodology, summarised clinical findings and study limitations. Where possible, correlation coefficients, MT mean and standard deviation were extracted to calculate effect sizes for meta-analyses.

### 2.4. Statistical Analysis

Descriptive statistics were calculated for demographic data, DMTs and steroid usage, and clinical disability measures. Key study findings and limitations were collated according to the MT technique used and the brain region.

When data were available from a sufficient number of studies, random-effects meta-analyses, with brain region as a nested factor, were performed to determine:

1. differences in MT metrics between patients with RRMS and healthy controls (significance level, α=0.05, *metafor* package in RStudio v1.3.1093).
2. putative relationships between clinical disability and MT metrics, in studies with reported correlation coefficients. Where the number of studies, *k*, was greater than two for a given brain region, follow-up sub-analyses were carried out to determine regional effect sizes, corrected for multiple comparisons (α=0.05/[1+ n of sub-analyses]). The Sidik-Jonkman method was used to assess between-study heterogeneity.

To assess longitudinal evolution of MT metrics in RRMS, longitudinal data (>1 time-point) were submitted to a mixed-model linear regression with mean MT as the dependent variable, time-point and brain region as fixed effects, and study as a random effect with within-study sub-grouping as a nested factor (e.g. active lesions versus reactivated lesions, placebo versus treatment groups; α=0.05; *lmer*, RStudio). Marginal means for each brain region were estimated (*ggeffects* R package). Follow-up sub-analyses were performed when *k*≥3 for a given brain region, with time-point as a fixed effect and study as a random effect, with sub-grouping as a nested factor (α=0.05/[1+ n of sub-analyses]).

### 2.5. Qualitative assessment

Longitudinal change in MT, the relationship between MT and treatment, its association with disability and the dependence on the MT metric used were qualitatively assessed, in addition to risk of bias.

## 3. Results

### 3.1. Systematic Online Literature Search Results

Initial online database searches yielded 6758 results. Following removal of duplicates, 3274 studies remained for abstract screening and abstracts were manually excluded according to outlined criteria (Table 1). Of the remaining 768 abstracts, full articles could not be retrieved for 42 abstracts and these were excluded.

**Table 1:**
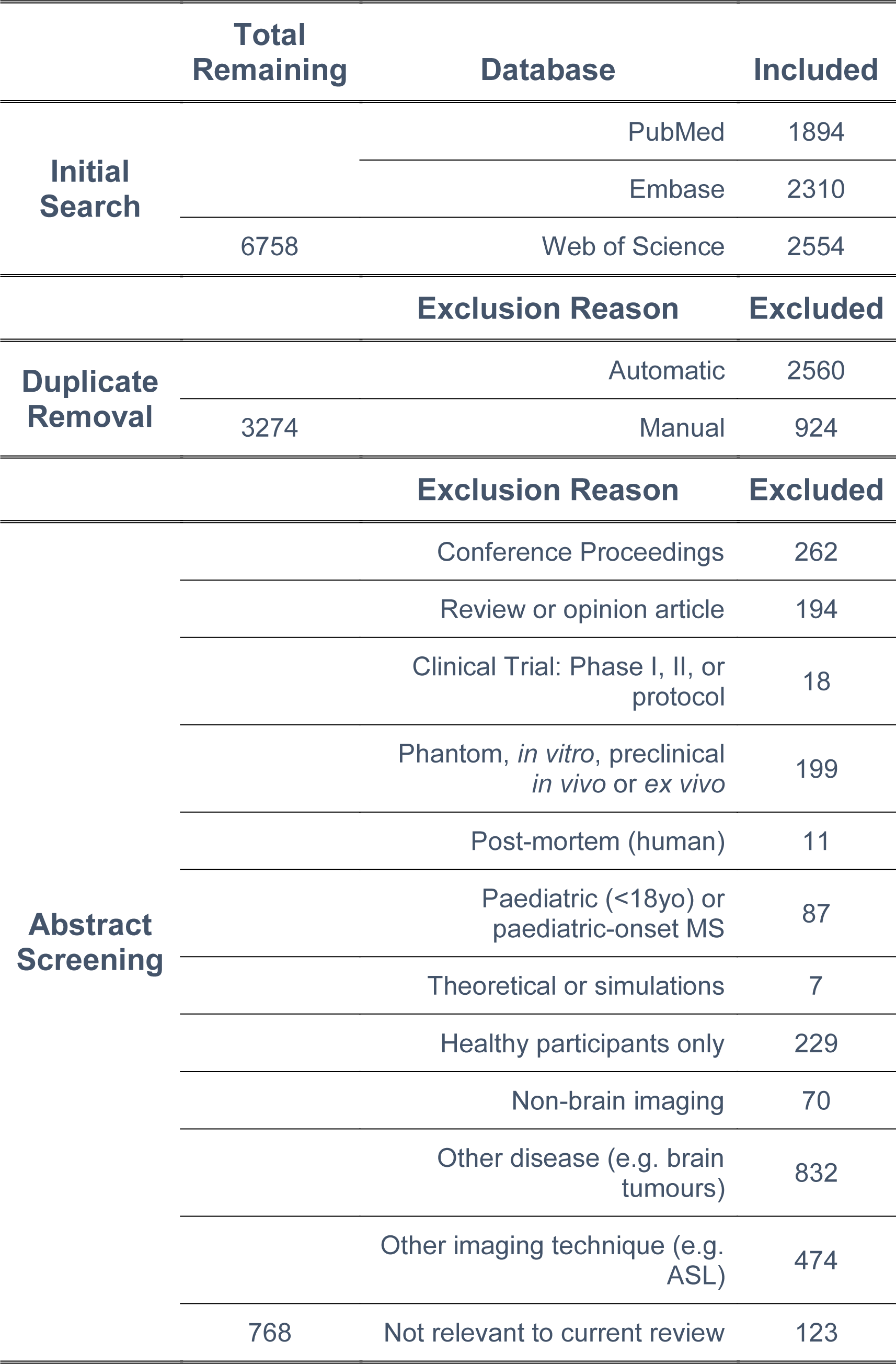
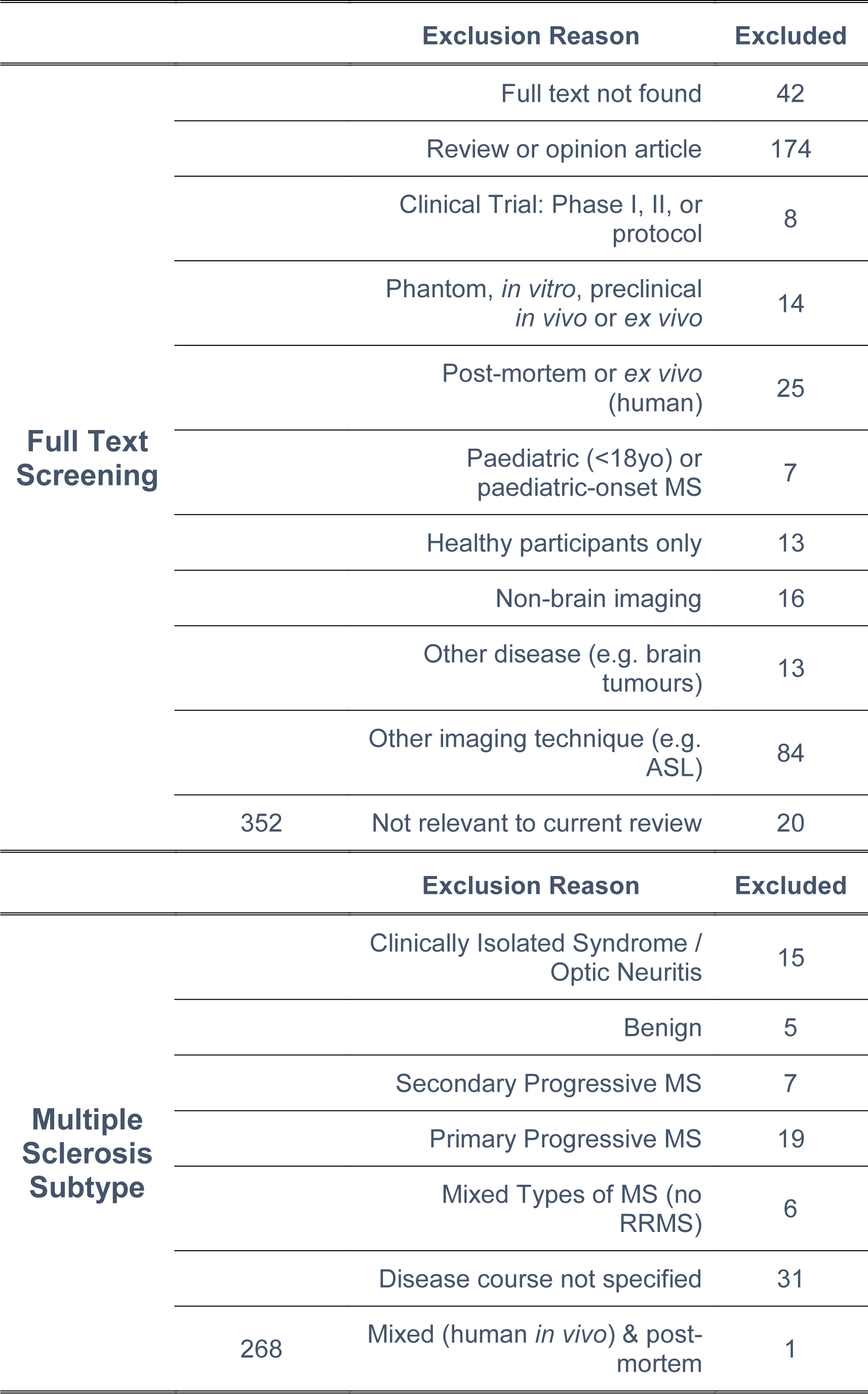
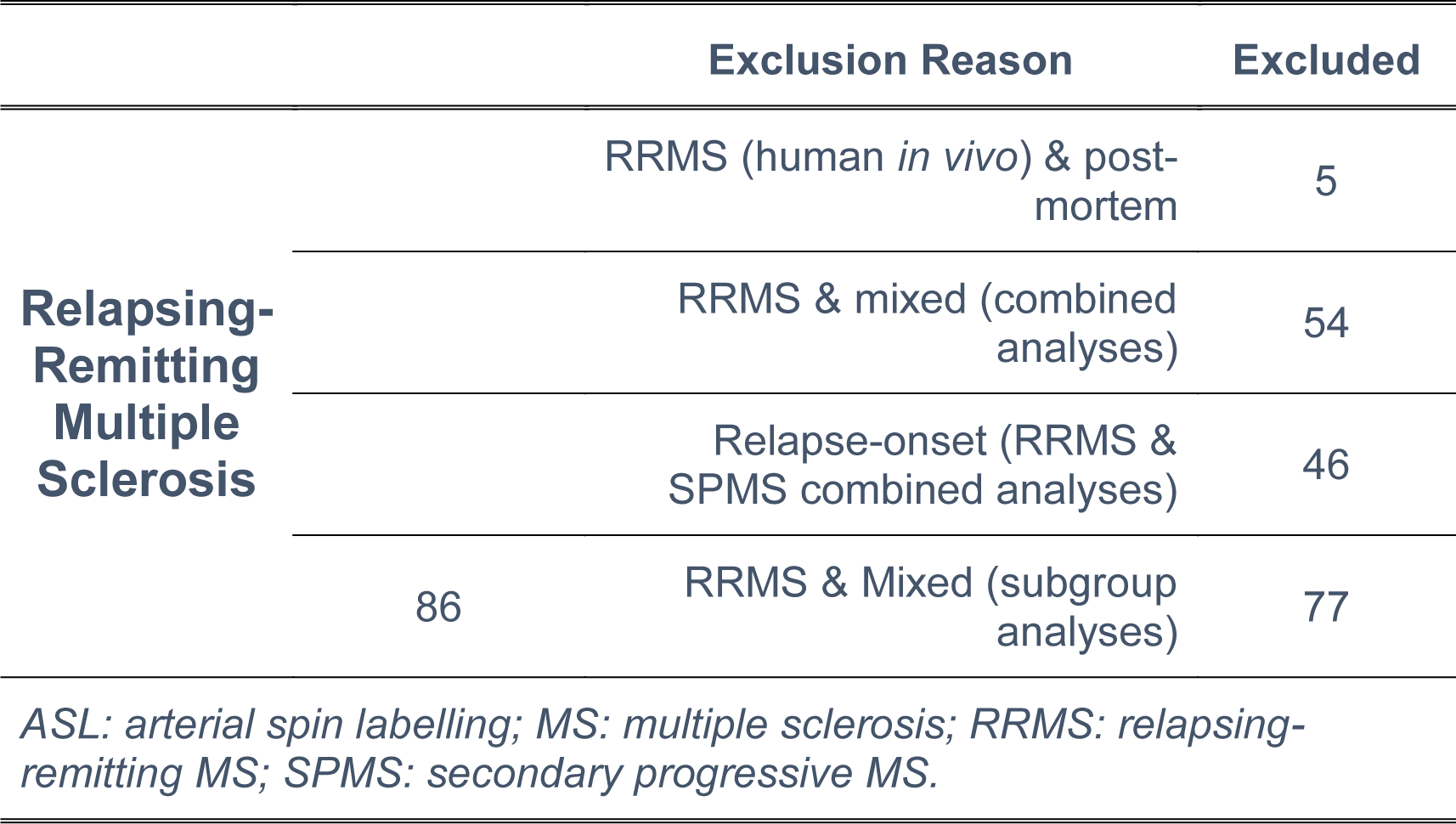
Overview of search process for systematic review.

Full text screening led to studies excluded for the following: review, commentary or opinion article (k=174), phase I/II clinical trial or protocols (k=8), non-human data (k=14), only post-mortem or ex vivo (human) data (k=25), paediatric subjects or paediatric-onset MS (k=7), healthy participants only (k=13), non-brain imaging (k=16), non-MS pathology (k=13), use of an imaging technique other than MTI (k=84), and not relevant to the current review (k=20).

As RRMS is the focus of this review, remaining studies (k=352) which only included participants with clinically isolated syndrome or optic neuritis (k=15), so-called benign multiple sclerosis (k=5), secondary progressive multiple sclerosis (SPMS, k=7), primary progressive multiple sclerosis (PPMS, k=19) or unspecified disease course (k=31) were excluded. Articles were rejected if participants had mixed disease courses other than RRMS (k=6), including any with additional post-mortem data (k=1).

The remaining selection (k=268) were refined by excluding studies in which RRMS patient data was combined before analysis with mixed multiple sclerosis subtype (k=54) or SPMS patient data (k=46), or included post-mortem data (k=5). Given the large number of articles remaining, studies which included multiple sclerosis subtypes other than RRMS but performed subgroup analyses were also excluded (k=77, references detailed in Supplementary Table 1 for mixed multiple sclerosis and Supplementary Table 2 for SPMS, combined with RRMS).

The final selection of articles (k=86, Supplementary Table 3) which form the foundations of this review thus only recruited participants with RRMS (and healthy controls, when included).

### 3.2. Sample Characteristics

#### 3.2.1. Sample Size

The median number of RRMS subjects recruited was 22 (range: 1^7, 30–33^ to 1512^34^), although the median number of patients with analysed MT data was lower (19, range: 1^7, 30–33^ to 858^34^, Supplementary Table 3). Fifty-seven studies (44%) included a healthy control group with a median of 14 (range: 2^33, 35^ to 56^36^) subjects recruited.^7, 30, 33, 35–88^

#### 3.2.2. Sex

The RRMS female-to-male ratio was two (median) at both recruitment (k=70/86 reported)^32, 36–38, 40–43, 45–49, 51, 52, 55–61, 63–65, 67, 70–72, 74–81, 83–86, 88, 89–107^ and analysis (k=61, Supplementary Table 3),^32, 34, 36–39, 41–49, 51–58, 60–65, 67, 68, 70–81, 83–87, 88, 89–98, 100, 101, 103–111^ compared with 1.43 for healthy controls (k=51)^a^.^30, 35–44, 46– 80, 83–85, 87, 88^

#### 3.2.3. Age

The mean age of people with RRMS^b^ was 37.15 years (5.63 SD, k=77, Table 2)^7, 30–32, 34–44, 46–49, 51–55, 57–75, 77–81, 83–89, 92–113^ and 35.70 years (4.90 SD, k=47) for controls.^35–43, 46–55, 57–68, 70–75, 77–79, 82–84, 87, 88^

**Table 2:**
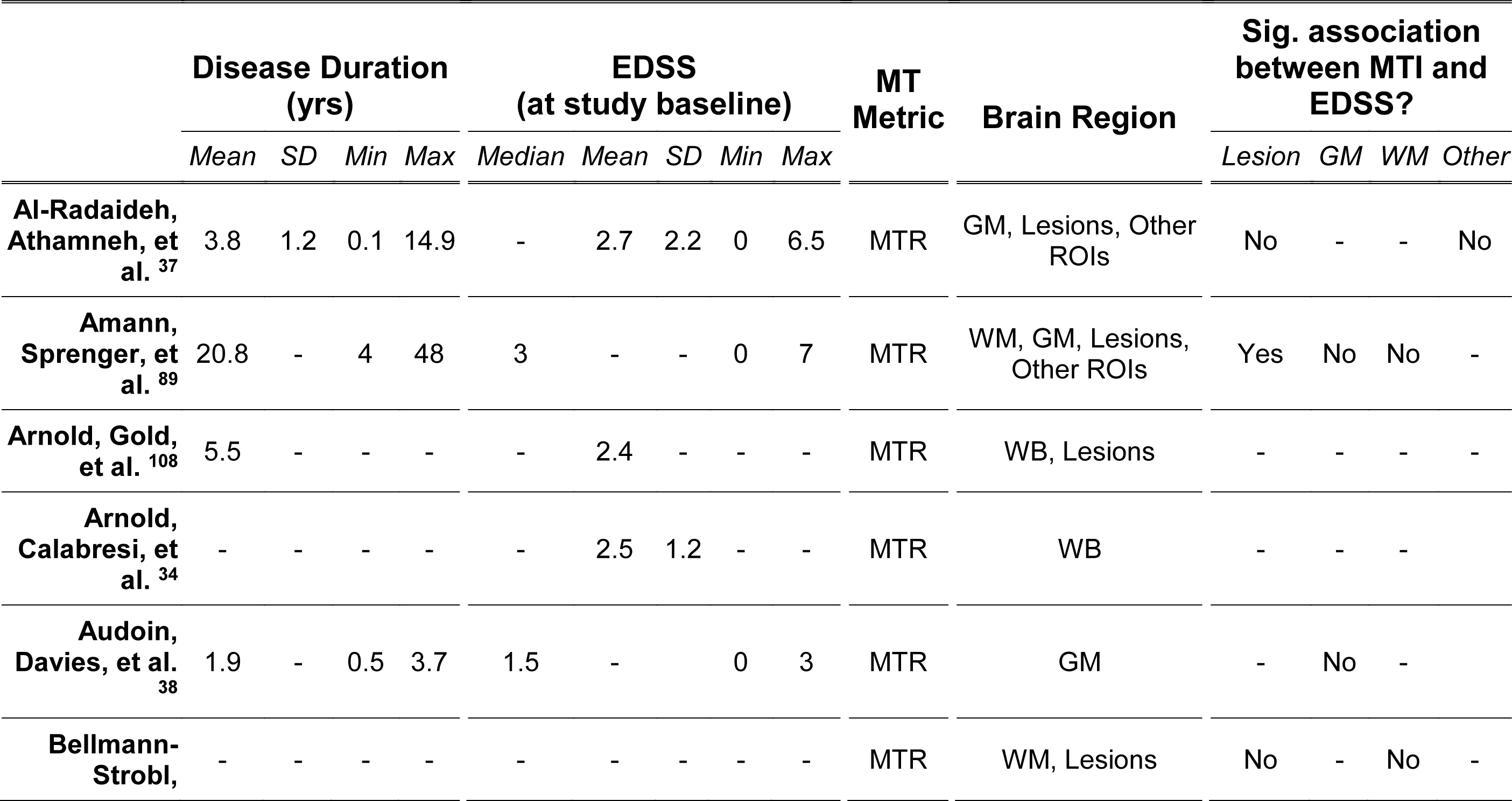
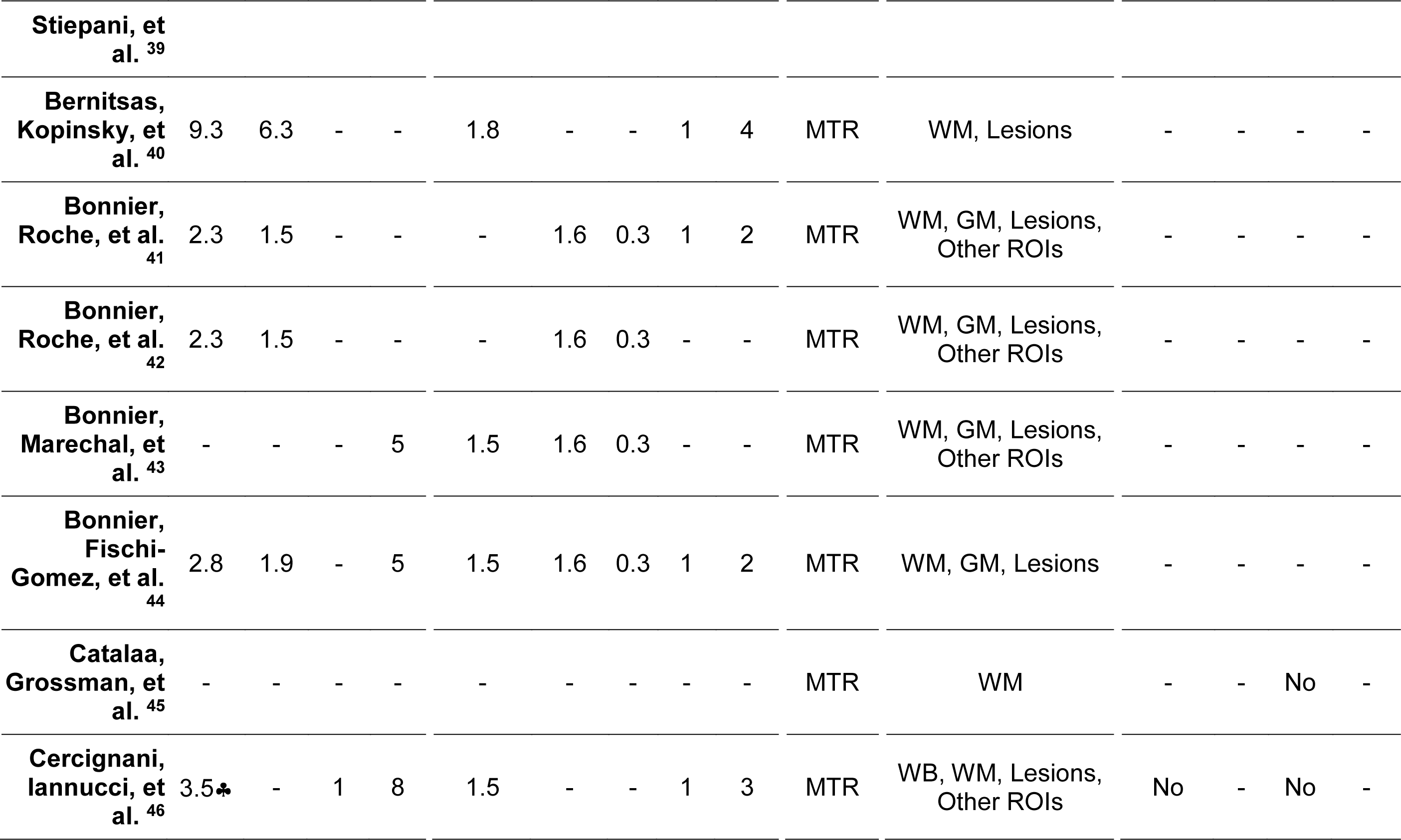
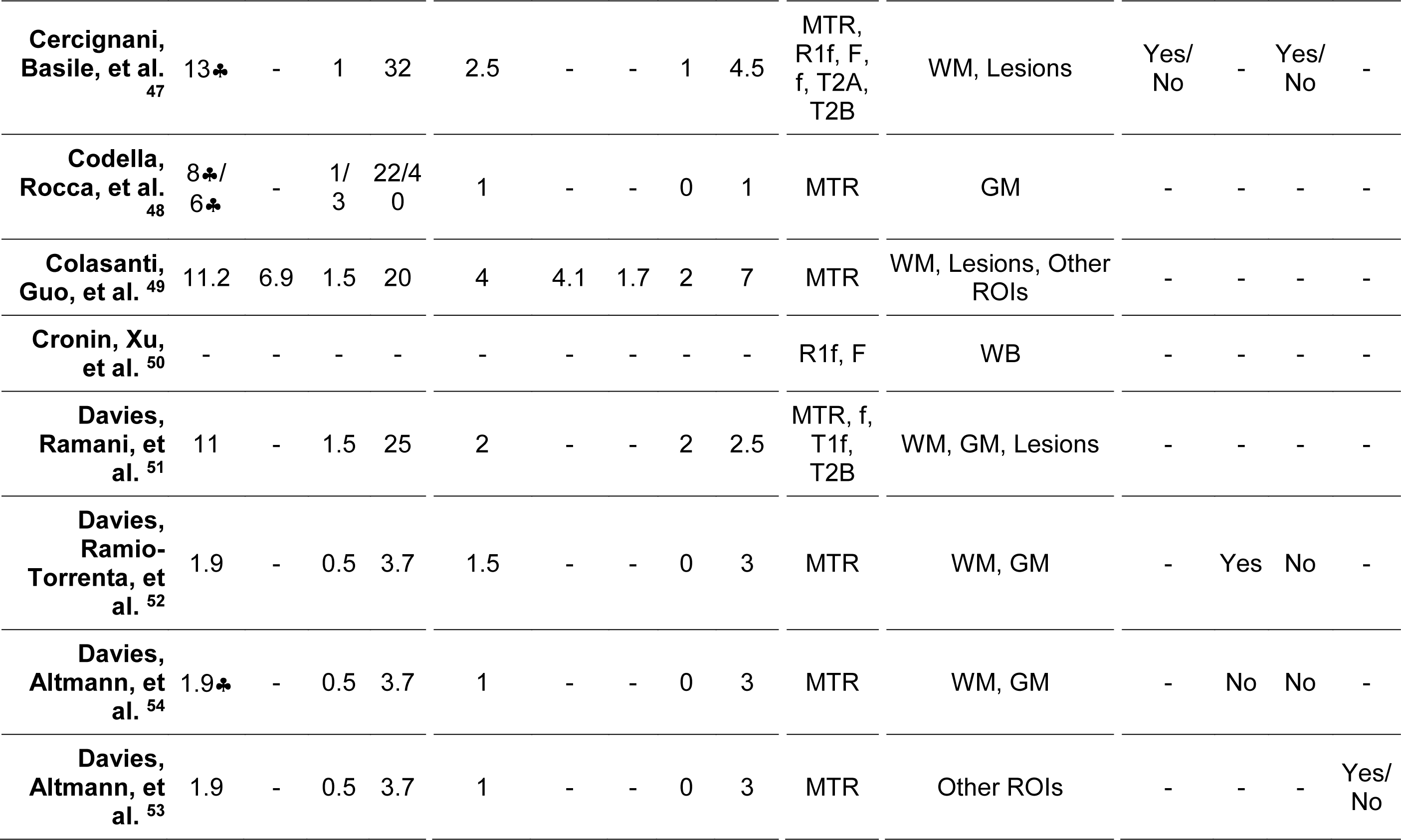
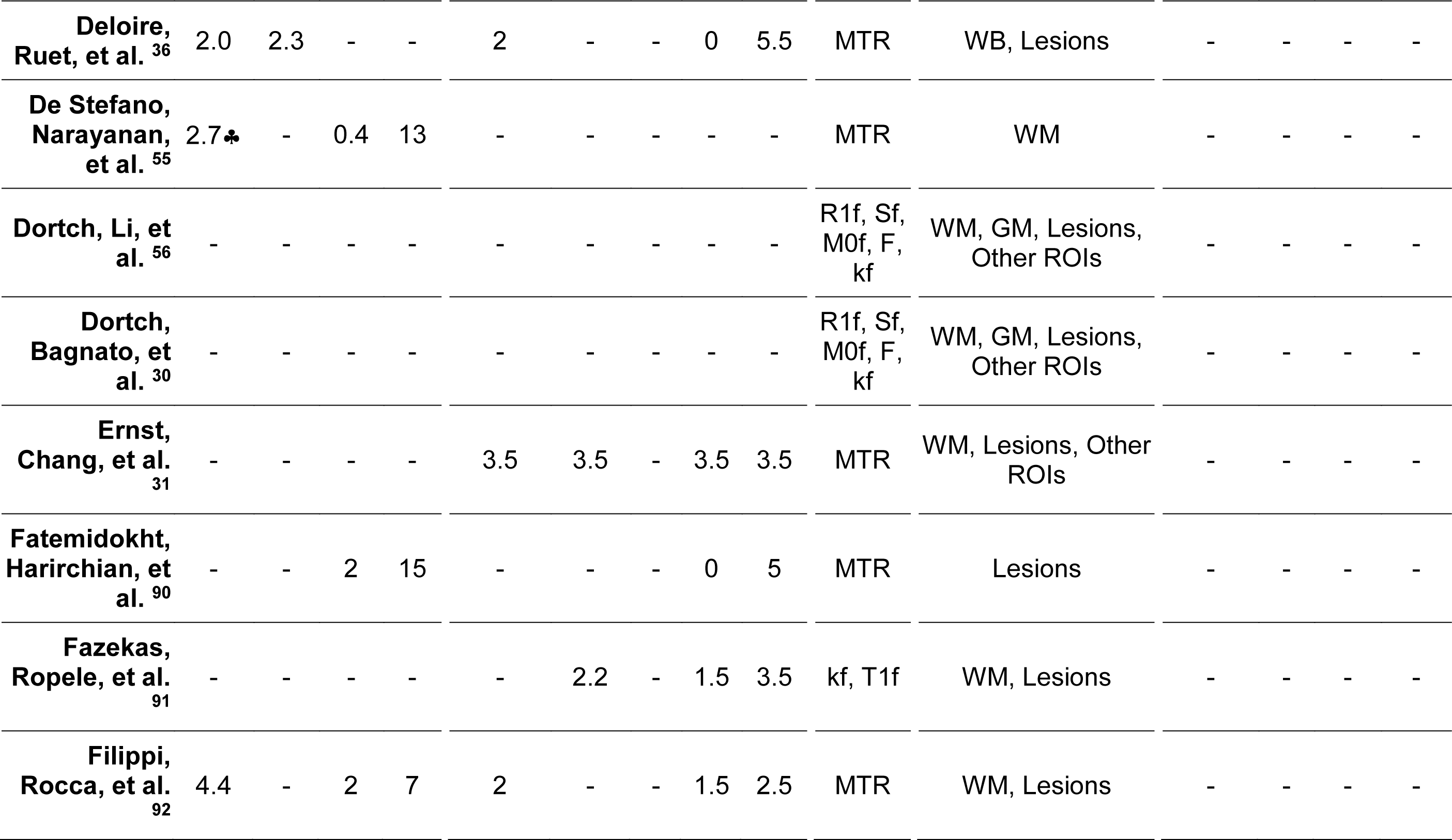
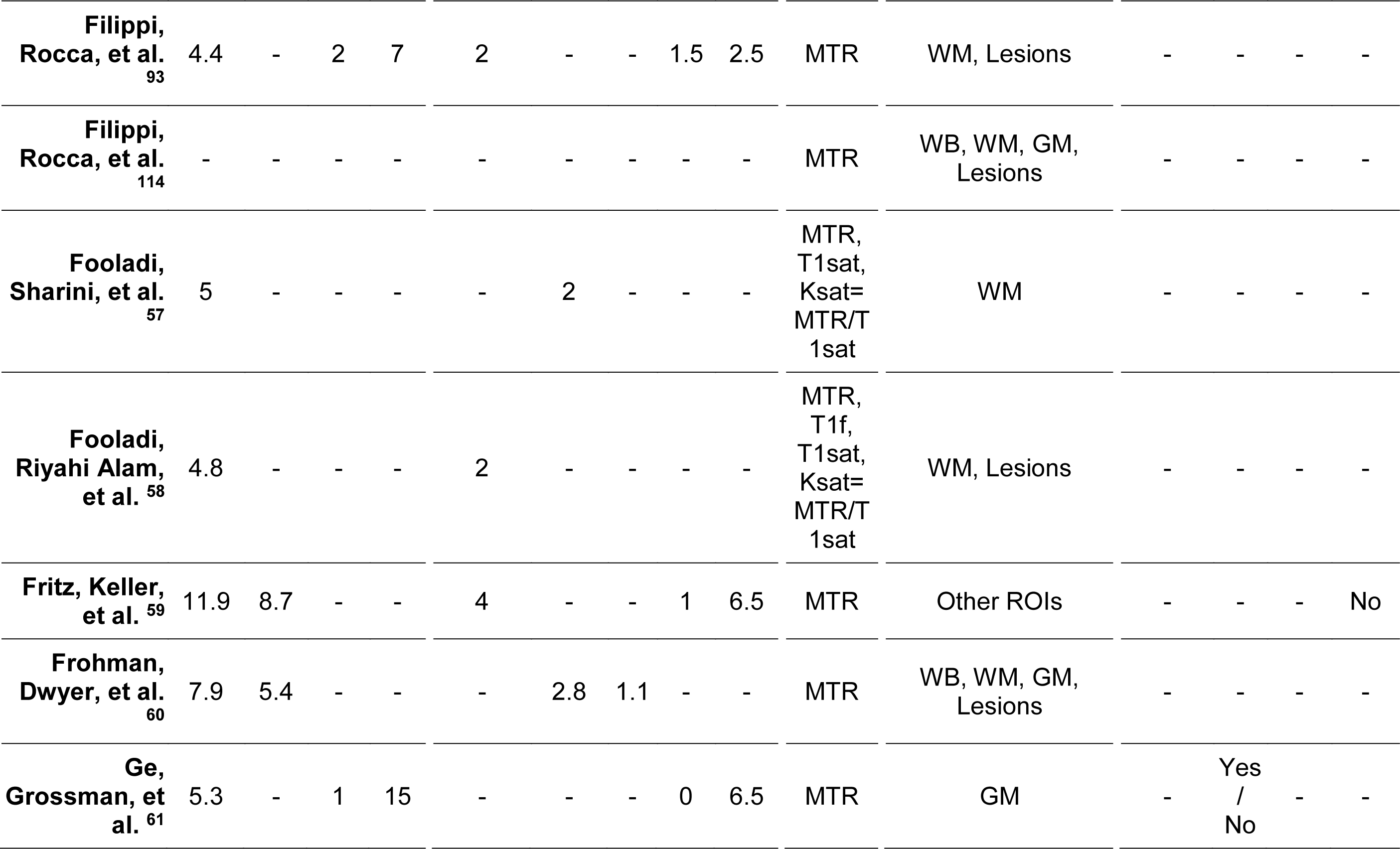
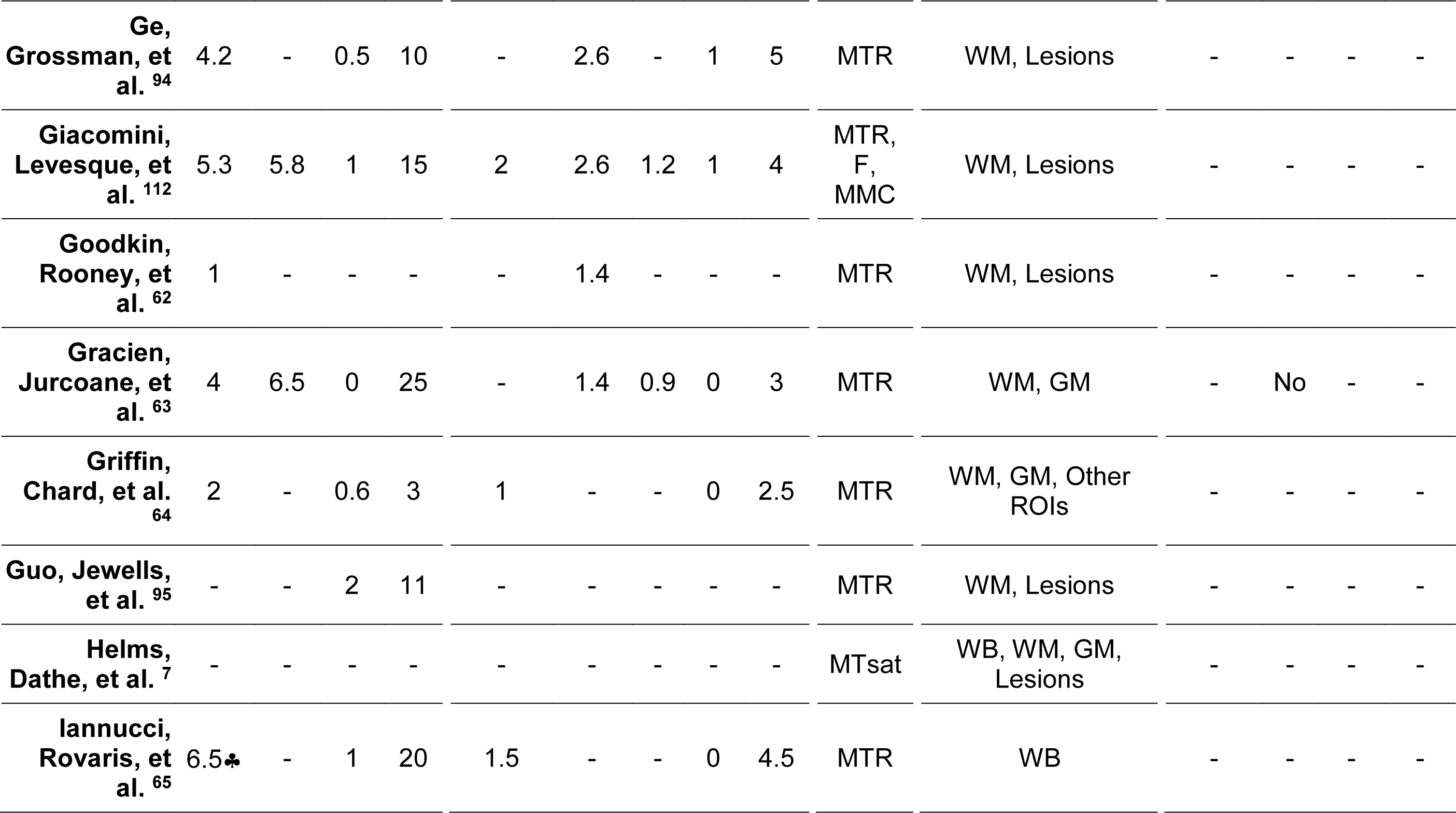
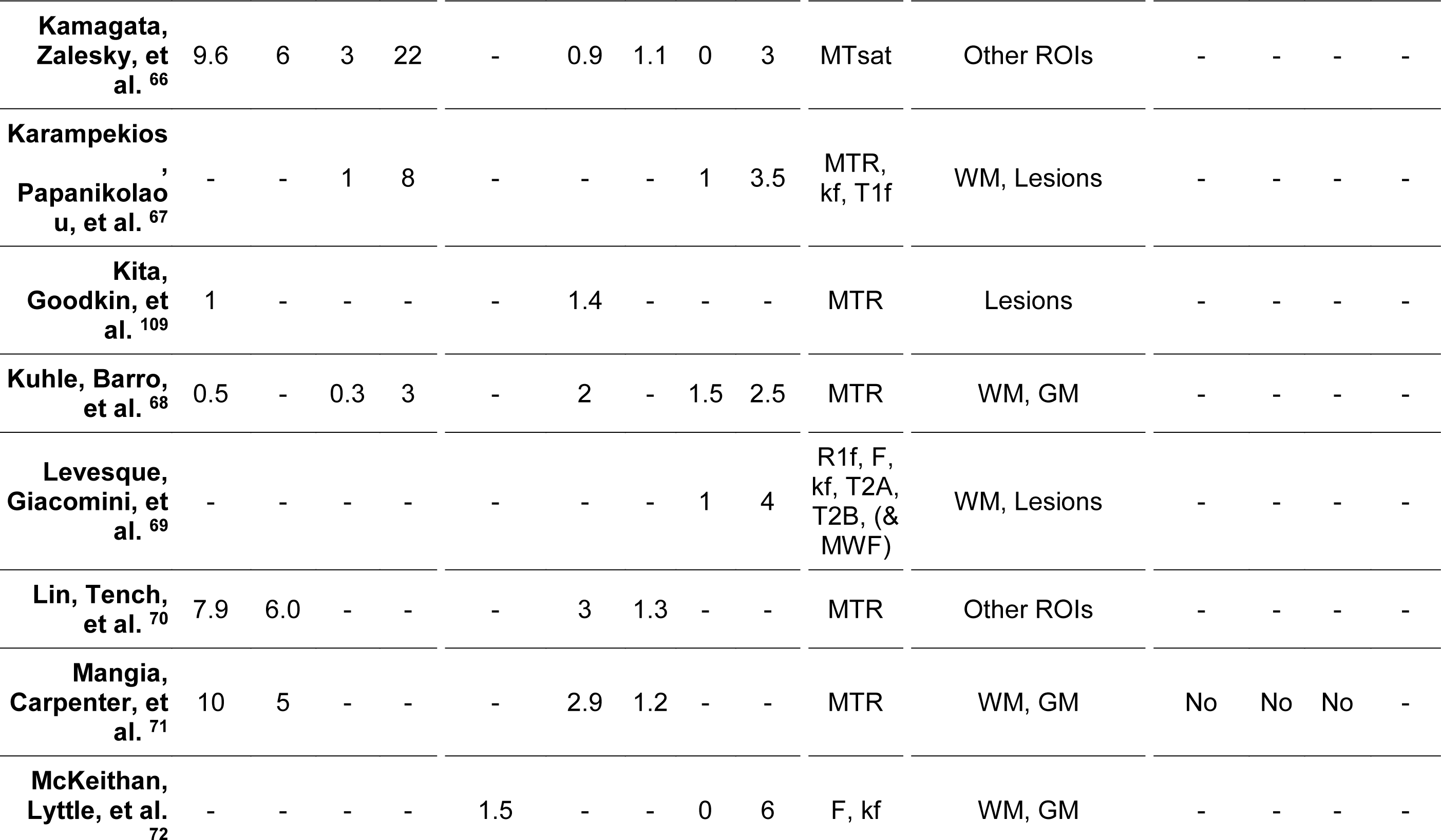
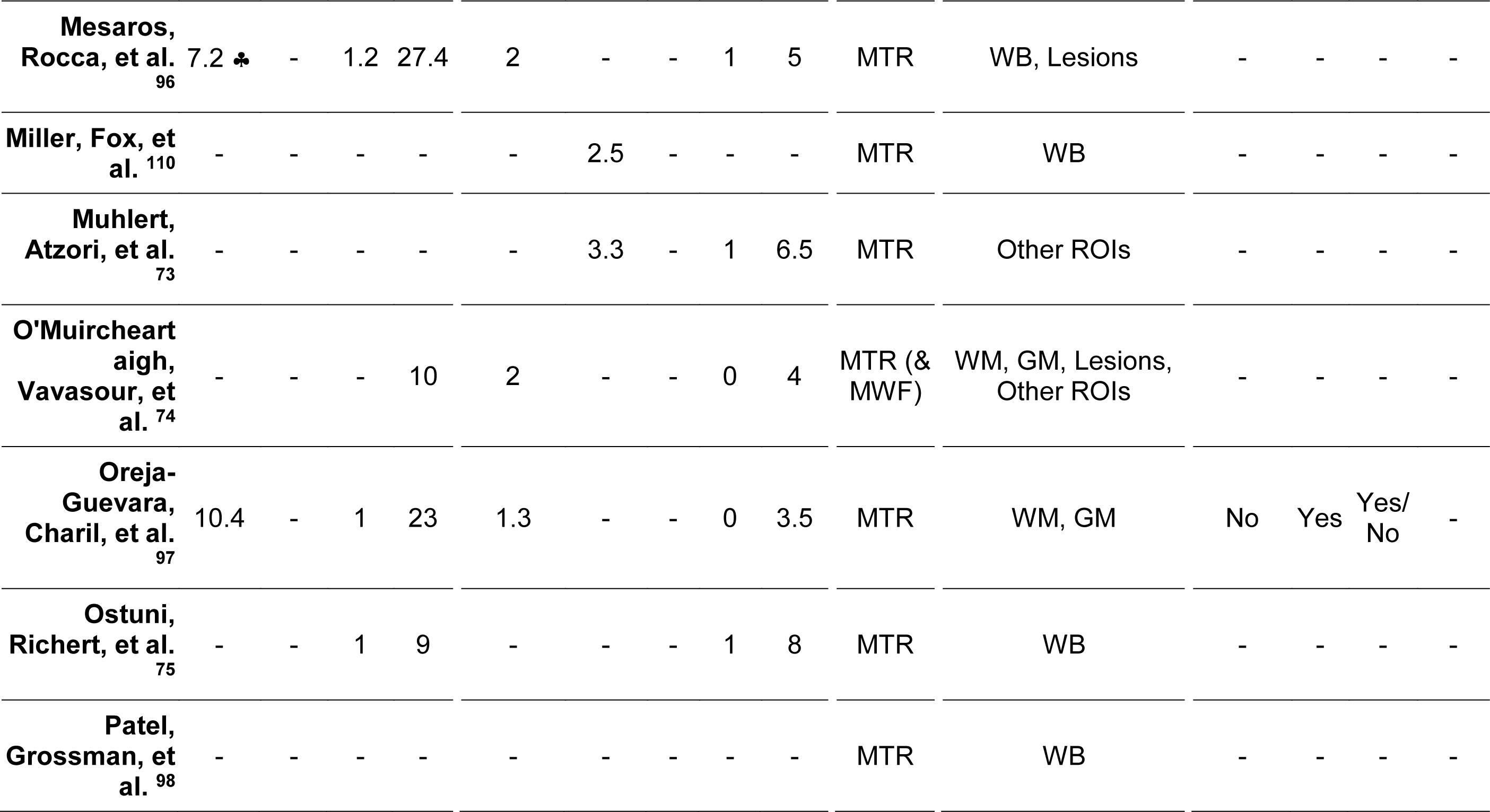
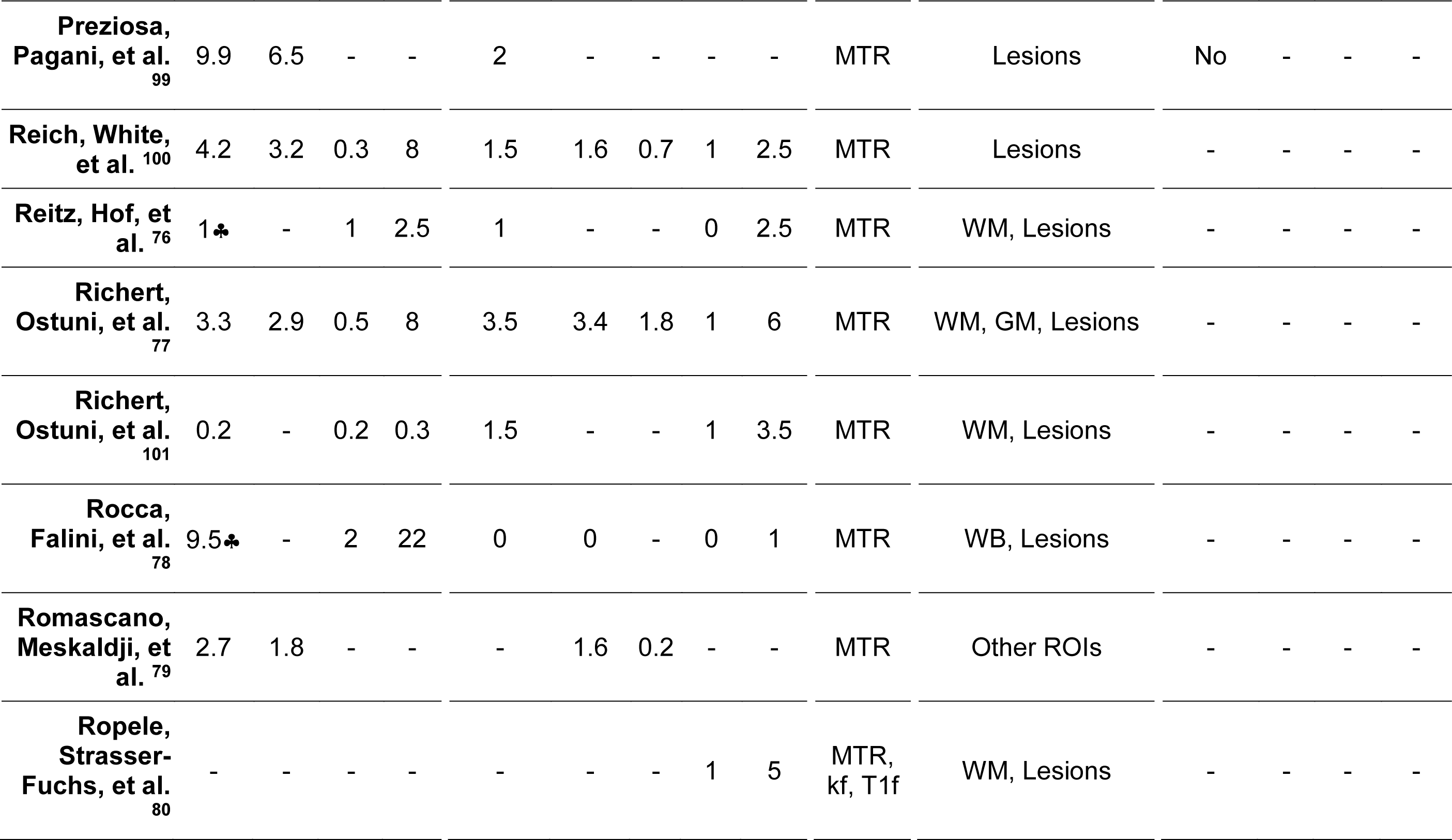
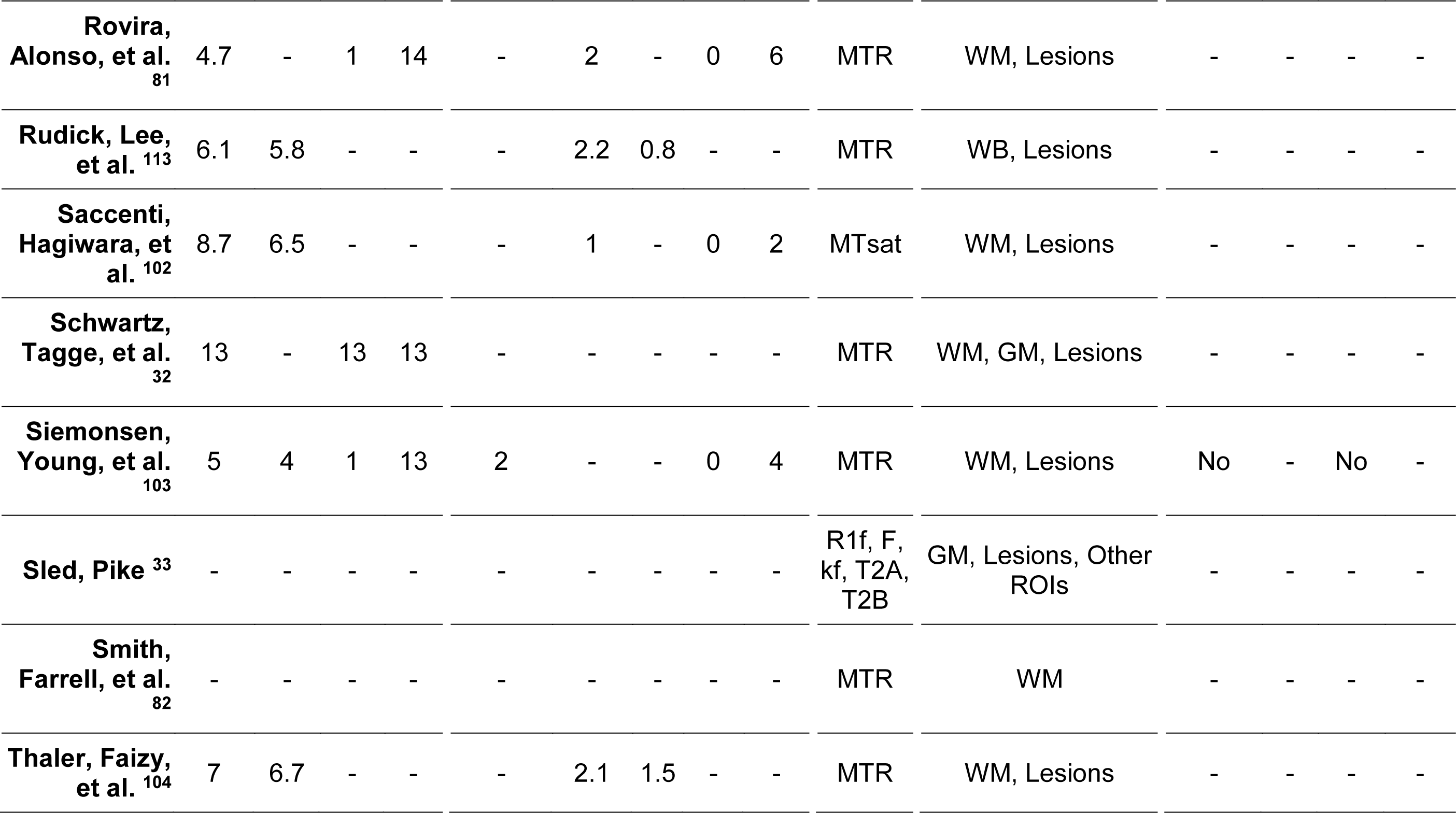
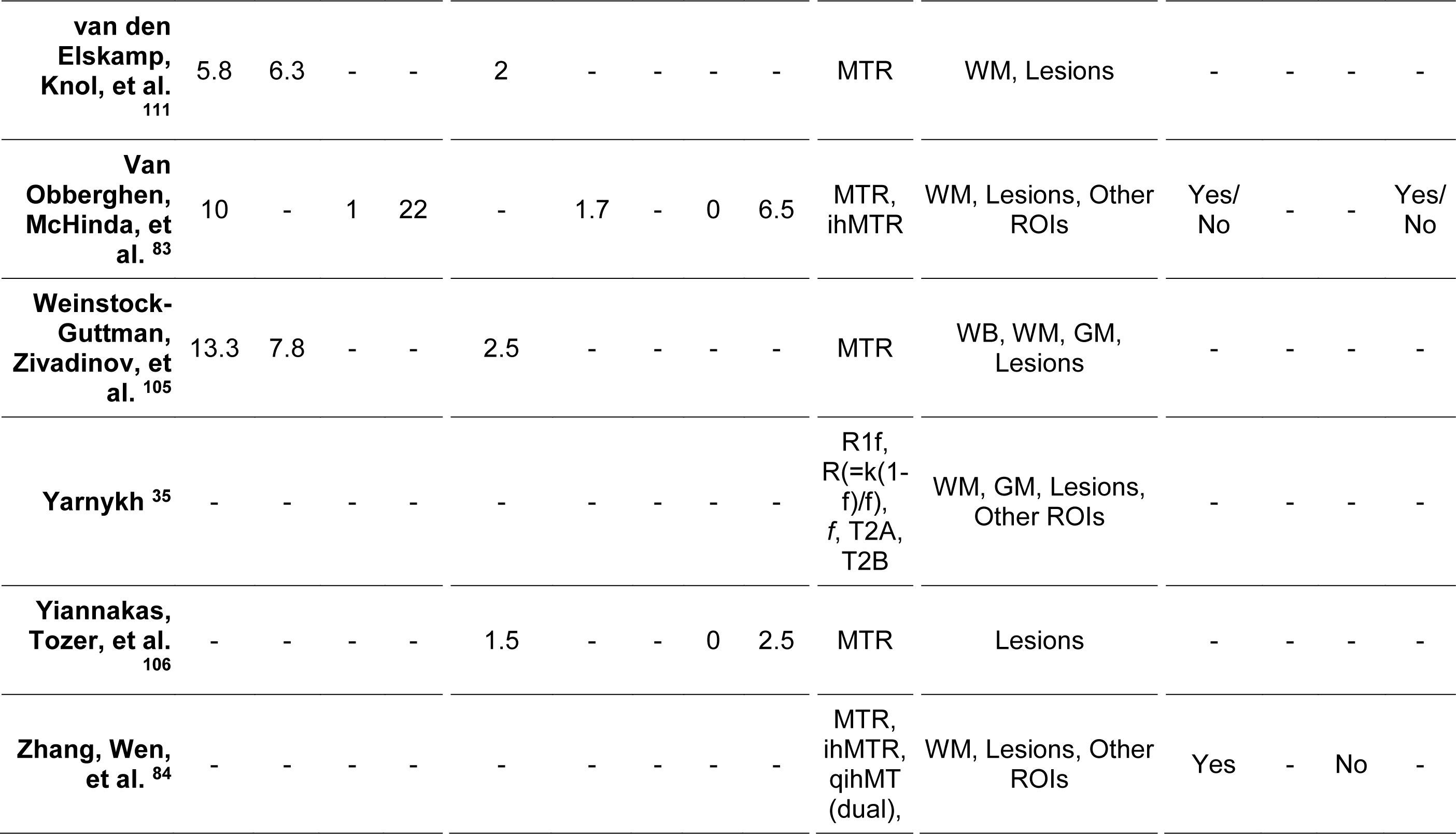
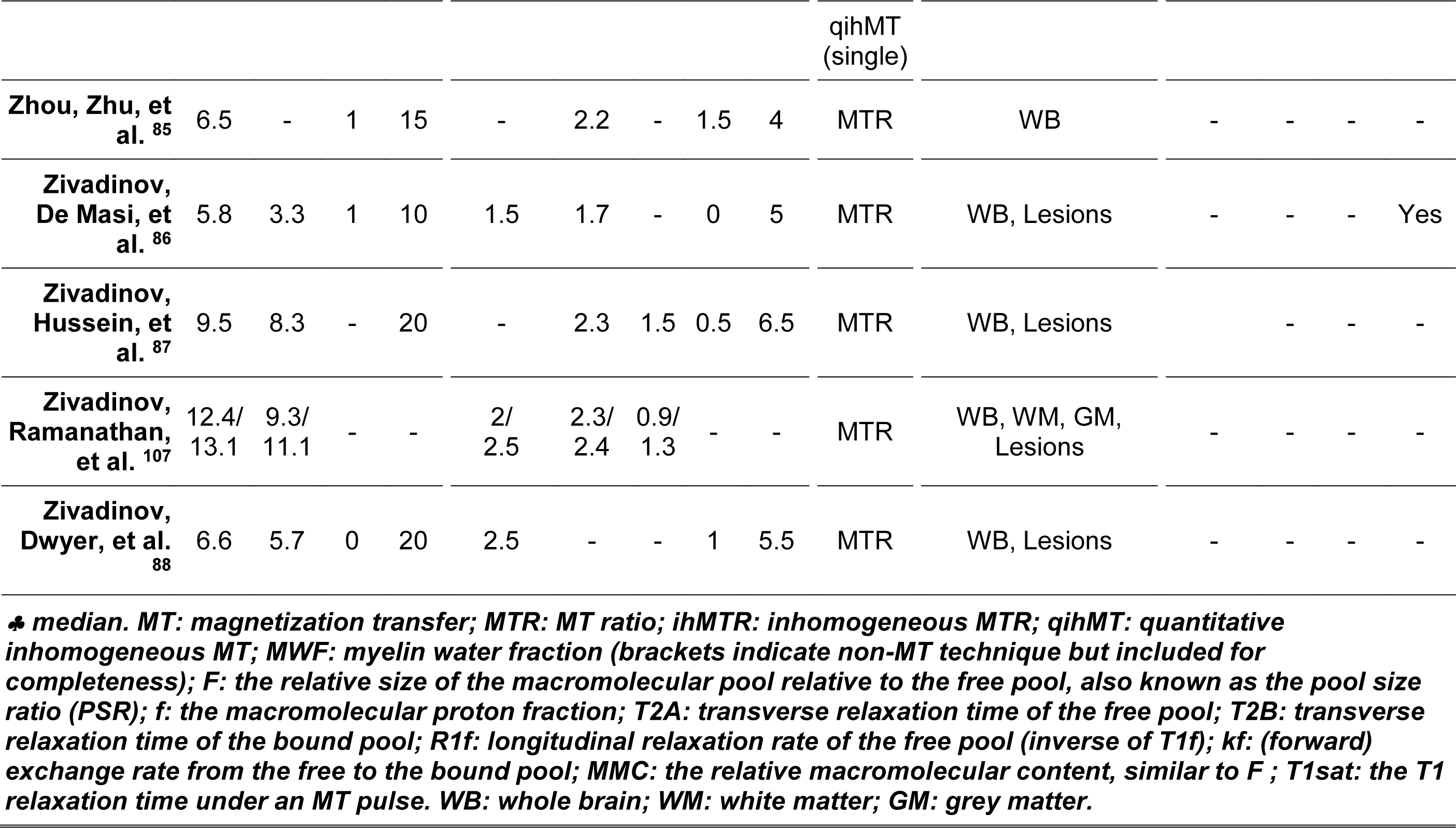
Overview of disease duration and clinical disability in relapsing-remitting multiple sclerosis cohorts with magnetisation transfer imaging.

#### 3.2.4. Nationality

The majority of studies were European (k=41/86, Supplementary Table 3)^7, 36, 38, 39, 41–44, 46–49, 51–54, 63–65, 67, 68, 70, 73, 76, 78–81, 83, 85, 86, 89, 91–93, 95, 97, 99, 103, 104, 106^ or North American (k=30),^30–32, 35, 40, 45, 50, 56, 59– 62, 69, 71, 72, 74, 75, 77, 82, 87, 88, 94, 98, 100, 101, 105, 107, 109, 112, 113^ with a minority of Asian^c^ (k=7)^37, 57, 58, 66, 84, 90, 102^ and international (k=8) studies (or >3 test centres).^33, 34, 55, 96, 108, 110, 111, 114^ The top three study locations were London (k=8),^49, 51–54, 64, 73, 106^ Milan (k=8),^46, 48, 65, 78, 92, 93, 97, 99^ and Lausanne (k=6).^41–44, 68, 79^

#### 3.2.5. Disease Duration

The mean disease duration across studies was 6.23 years (4.19 SD, k=50/86 reported as mean, Table 2)^32, 36–38, 40–42, 44, 49, 51–53, 57–64, 66, 68, 70, 71, 77, 79, 81, 83, 85–89, 92– 94, 97, 99–105, 107–109, 111–113^ with range 0.20101 to 20.80 years89.

#### 3.2.6. Clinical Disability

The majority of studies (k=73/86) used the Expanded Disability Status Scale (EDSS) as a measure of disability (Table 2) with median^d^ baseline score of 1.5 (k=64).^31, 34, 36–38, 40–44, 46–49, 51–54, 57–60, 62–66, 68, 70–74, 76–79, 81, 83, 85–89, 92–94, 96, 97, 99–113^ Additional clinical correlates included the Multiple Sclerosis Functional Composite (MSFC, k=11)^36, 38, 39, 41, 42, 52–54, 71, 79, 113^ or its subcomponents, i.e. the Paced Auditory Serial Addition Test (PASAT), 9-Hole Peg Test (9HPT) or the Timed 25-Foot Walk (T25FW, k=5),^43, 59, 70, 72, 86^ the Symbol-Digit Modalities Test (SDMT), Stroop test, Weschler Abbreviated Scale of Intelligence, Adult Memory and Information Processing Battery, Hospital Anxiety and Depression Scale,^73^ Hamilton Depression and Anxiety Rating Scales, Mini- Mental State Examination, and the Standard Raven Progressive Matrices.^86^

#### 3.2.7. Disease Modifying Therapies (DMTs) and Steroid-Usage

Intra-study and inter-study heterogeneity were apparent in treatment with DMTs and steroids (Table 3 and Supplementary Table 4 for summaries; Supplementary Table 3 for detailed descriptions). Homogeneous DMTs were prescribed across the cohort in eleven studies (Supplementary Table 4); comprising fingolimod,^40^ dimethyl fumarate,^32, 104^ subcutaneous interferon (IfN)-β1a,^39, 88^ or IfN-β1b,^31, 77, 101^ intramuscular IfN-β1a,^107, 109^ and subcutaneous glatiramer acetate.^87^ Patients in four further studies were either untreated or received homogeneous DMTs which were IfN-α^98^ IfN-β,^53, 54^ and glatiramer acetate.^69^

**Table 3:** Overview of use of DMTs for patients with relapsing-remitting multiple sclerosis in studies using magnetisation transfer imaging. Studies may be duplicated where treatments were heterogeneous. Study-specific details are given in Supplementary Table 3.

| DMTs | k | % | Citation |
| --- | --- | --- | --- |
| dimethyl fumarate | 4 | 4.7% | 32,63,76,104 |
| dimethyl fumarate (delayed release) | 2 | 2.3% | 108,110 |
| fingolimod | 10 | 11.6% | 40-44,63,68,76,79,99 |
| natalizumab | 5 | 5.8% | 49,63,76,99,112 |
| glatiramer acetate | 9 | 10.5% | 47,63,68,69,76,87,100,110,112 |
| Interferon-β (1a) | 13 | 15.1% | 39,68,74,76,88,91,98,100,107,109,111-113 |
| Interferon-β (1b) / Betaferon | 5 | 5.8% | 31,68,77,101,112 |
| interferon beta (unspecified) | 8 | 9.3% | 41-44,47,53,54,79 |
| pegylated interferon 1a | 1 | 1.2% | 34 |
| laquinomod | 1 | 1.2% | 114 |
| ocrelizumab | 1 | 1.2% | 74 |
| placebo | 8 | 9.3% | 36,49,58,80,83,89,113 |
| Steroids | k | % | Citation |
| methylprednidolone | 2 | 2.3% | 31,101 |
| Unspecified | 2 | 2.3% | 94,98 |
| None (for indicated time period) | 26 | 30.2% | 32,37,38,40,43,44,46-49,55,59,60,62,64,73,78,81,85,86,89,92,93,97,105,109 |
| Data missing | 56 | 65.1% | 7,30,33-36,39,41,42,45,50-54,56-58,61,63,65-72,74-77,79,80,82-84,87,88,90,91,95,96,99,100,102-104,106-108,110-114 |

Patients in five studies were treatment-naïve (and not receiving steroid treatment for a minimum of fourteen days prior to imaging),^45, 52, 92–94^ and only the placebo-arm of a clinical trial was included in one study.^96^ Eleven studies allowed steroid treatment for relapses or did not specify usage, but were otherwise treatment- naïve.^38, 46, 48, 55, 60, 62, 64, 78, 81, 86, 97^ Many studies did not report DMT or steroid usage (k=28 and k=56, Supplementary Table 4 and Table 3, respectively) or did not specify DMTs (k=5).^36, 58, 80, 83, 89^ However, studies that reported steroid usage typically had a washout period of at least ten days before MR imaging took place.

### 3.3. MTI Acquisition Protocol Parameters

MTI protocols varied across studies, and parameters were often unreported.

#### 3.3.1. Magnetic field strength

MTI was mainly performed at 1.5T7,^31, 33, 36, 38, 39, 45, 46, 48, 51–54, 57, 58, 60– 62, 64, 65, 67, 69, 70, 75, 77, 78, 80, 81, 83, 85–87, 89, 91–98, 100, 101, 105–107, 109, 111, 112, 114^ (k=50, Figure 1A).

Recent studies were acquired at 7T,72 3T (k=29),^30, 32, 35, 37, 40– 44, 47, 49, 50, 56, 59, 63, 66, 68, 73, 74, 76, 79, 82, 84, 88, 90, 99, 102–104^ both 3T and 1.5T,34 or 4T.71 Field strength was occasionally unreported (k=4), although 1.5T may be assumed in such cases due to publication dates or multi-centre approaches.^55, 108, 110, 113^

#### 3.3.2. MTI Sequence

##### 3.3.2.1. Pulse sequence

In the majority of MTI protocols (k=60), gradient echo (GRE) was used, either as a 2D (k=13),^46, 48, 51, 65, 70, 78, 81, 85, 92, 93, 96, 97, 111^ 3D (k=27), ^7, 32, 34, 35, 37, 45, 47, 49, 57–63, 66, 67, 72–74, 82, 84, 86, 90, 94, 98, 99, 102, 109, 113^ both 2D and 3D (k=1),89 or unspecified (k=16)33,39-^44, 55, 68, 69, 76, 79, 103, 104, 112, 114^ GRE sequence. Eight studies used 2D spin echo (SE, k=8)^38, 52–54, 64, 83, 100, 106^ and one study employed both SE and GRE.^50^ The pulse sequence was not described in six studies.^31, 36, 87, 88, 108, 110^

##### 3.3.2.2. Image contrast: TR, TE and excitation flip angle

Proton density (PD)-weighting (with and without an MT pulse) was typically used for MTI. Nevertheless, some studies adopted T1-weighting^77, 99–101, 104^ or T2*-weighting.^41– 44, 68, 79^ The TR and TE varied accordingly with the intended weighting and sequence type. For example, TR ranged from 2.67 ms for a MT-sensitized balanced steady- state free precession sequence (bSSFP) at 1.5T^89^ to 3000 ms for a 2D pulsed inhomogeneous MT HASTE (Half-fourier Acquisition Single-shot Turbo spin-Echo),^83^ with a median TR of 65.50 ms (k=74/86). The median TE was 11.70 ms (k=73, range: 1.23 ms^41–44, 68, 79^ to 90 ms^38, 52–54, 64^). Some studies did not, however, report TR (k=12)^30, 31, 39, 50, 56, 58, 71, 72, 87, 91, 108, 110^ or TE (k=13). ^31, 33, 39, 58, 69, 71, 72, 87, 91, 102, 108, 110, 112^ The excitation flip angle ranged from 3° for a 3D FLASH (fast low-angle shot) acquisition^67^ to 90° for 3D selective inversion recovery (SIR)-turbo SE & 3D SIR-EPI (echo planar imaging),^50^ T1-weighted 2D SE^100^ and PD-weighted MT sequences,^88^ with a median of 15° (k=55). Many studies (k=31) did not report the excitation flip angle.^30, 31, 38, 39, 41–44, 52–56, 59, 64, 68, 71, 72, 75, 77, 83, 87, 90, 91, 95, 101, 105–108, 110^

##### 3.3.2.3. Voxel Size, Slice Thickness and Number of Slices

The median in-plane voxel size was 1.0mm by 1.0mm (k=73 & k=66/86 respectively, range: 0.7mm^51^ to 2.2mm^50, 56, 82^). The median slice thickness was 4.0 mm (k=73/86, range: 1mm49,73 to 9mm83 7,^30, 32–38, 40–46, 49–66, 68–70, 72–79, 81–83, 85, 86, 88–91, 93, 95–107, 109, 111–114^).

The median number of slices acquired was 28 (k=61, range: 130,33,45,56,69,112 to 192102 ^30, 32, 33, 35, 36, 38, 40, 42–46, 48–50, 52–58, 60, 63, 64, 66, 68–80, 82–86, 89–91, 93, 96–99, 101–107, 112, 114^) with resulting coverage of 126mm (median, k=55, range: 5mm30,45,56 to 280mm60).

#### 3.3.3. MT Pulse Characteristics

##### 3.3.3.1. Radiofrequency Pulse Frequency

The vast majority of studies achieved selective saturation of the ‘bound’ pool with a radiofrequency pulse at an offset or multiple offsets from the water proton frequency (Figure 1B). When a single offset was used (k=59/86), the median frequency was 1500 Hz (range: 600 Hz^101^ to 7000 Hz^83^). When multiple offset frequencies were considered to permit quantitative model construction or due to inter-centre variability, the range was wider (100 Hz to 80 kHz, k=7).^33, 35, 45, 47, 69, 112, 114^

**Figure 1:**
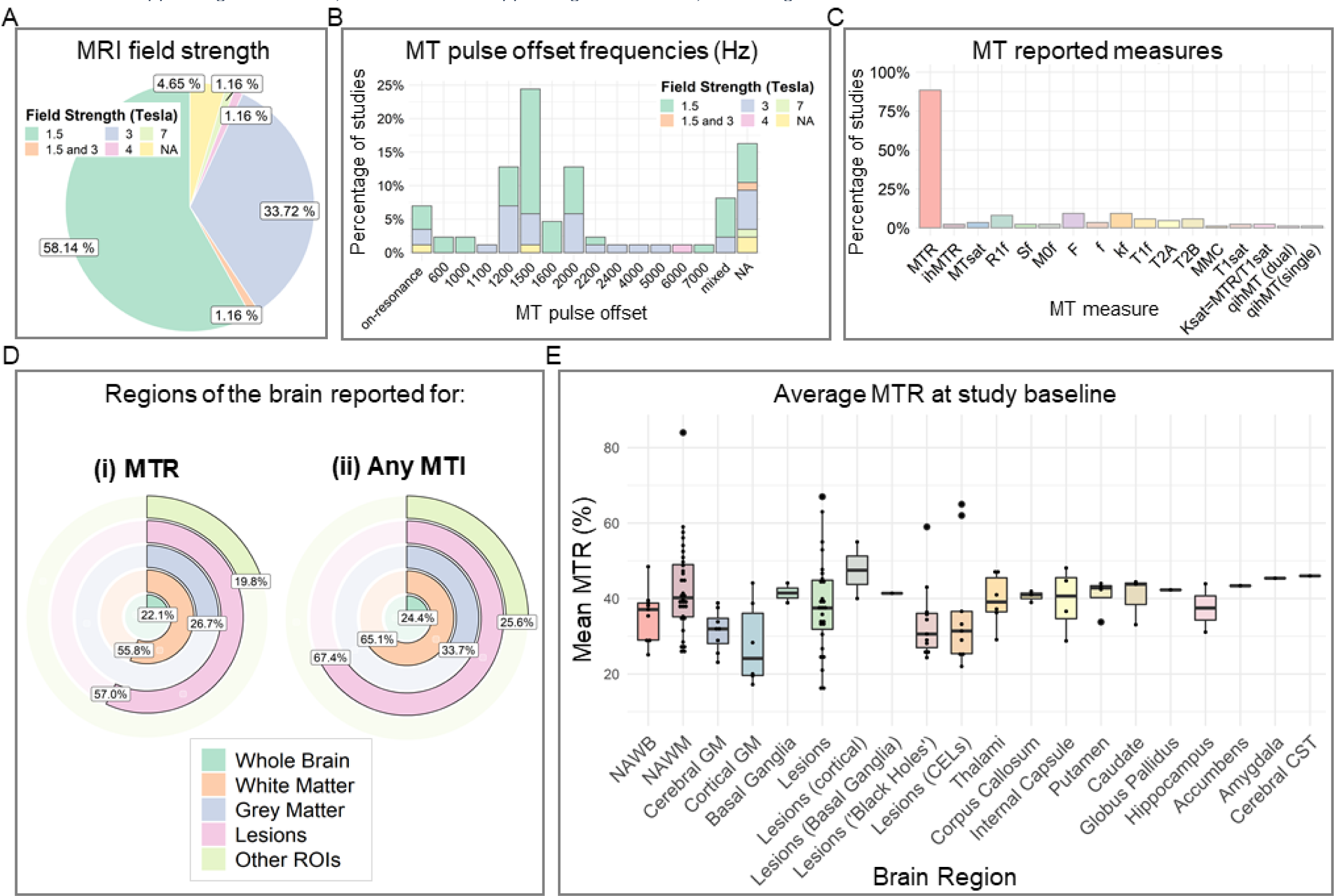
MRI characteristics of studies which used magnetisation transfer imaging in relapsing-remitting multiple sclerosis (k=86). CELs: contrast- enhancing lesions; CST: corticospinal tract; GM: grey matter; MMC: macromolecular content; MT: magnetisation transfer; MTR: MT ratio; ihMTR: inhomogeneous MTR; MTsat: MT saturation; qihMT: quantitative inhomogeneous MT; NAWB: normal- appearing whole brain; NAWM: normal-appearing white matter; ROIs: region of interest..

Alternative approaches included SIR with a low-power on-resonance pulse^30, 50^ and FastPACE two-point T1 mapping with a 1-2-1 binomial on-resonance pulse.^80, 91^ SIR aims to invert the ‘free’ water while leaving the ‘bound’ pool relatively unaffected.

Quantitative SIR MTI parameters are estimated from the bi-exponential recovery of the ‘free’ water signal, sampled at various inversion times. Two studies used on-resonance pulse MTI but did not report further details^55, 86^ and fourteen studies did not report the offset frequency.^31,34,37,56,68,72,79,87,94,99,100,106,108,110^

##### 3.3.3.2. Flip angle of MT Pulse

Amongst studies that reported the MT pulse as a flip angle, the median angle was 500° (k=34) with range 200°^43^ to 1430°.^38, 52, 53, 64^ When only pulse peak amplitude was reported, the range was 3.4μT^92, 93, 97^ to 23.6μT,^80^ with a median of 7μT (k=15). Four studies used multiple pulse energies^33, 35, 69, 112^ and a number of studies (k=33) did not report the pulse flip angle or power.^30, 31, 34, 36, 37, 47, 50, 51, 53, 56, 59, 60, 68, 71, 72, 75, 79, 81, 86–88, 90, 94, 99–101, 105–108, 110, 113, 114^

##### 3.3.3.3. Shape of MT Pulse

The radiofrequency pulse shape was generally Gaussian (k=28),^7, 32, 46– 49, 62, 63, 65–67, 70, 73, 78, 81, 85, 86, 89, 92, 93, 96, 97, 102–104, 109, 113^ although Sinc (k=6),40,45,61,94,95,98 Sinc-Gaussian (k=5),^35, 59, 74, 82^ Fermi (k=4),^57, 58, 84, 90^ 1-2-1 binomial (k=2),^80, 91^ and hyperbolic secant pulses (k=1)^71^ were also used. Forty studies did not specify pulse shape.

##### 3.3.3.4. Pulse Duration

The radiofrequency pulse duration was related to the type of MT sequence and ranged from 0.08 ms for MT-sensitized bSSFP^89^ to 700 ms for 2D-pulsed-ihMT HASTE sequence.^83^ Median MT pulse duration was 10.15 ms (k=61).

### 3.4. Quantitative Measures of Magnetisation Transfer

#### 3.4.1. Metrics used

The most frequently used quantitative MT metric was MTR (k=75, Figure 1C and Table 2).^31, 32, 34, 36–49, 51–55, 57–65, 67, 68, 70, 71, 73–90, 92–101, 103–114^ A small number of studies used MTsat (k=3),^7, 66, 102^ ihMTR or quantitative ihMT (k=2),^83, 84^ or qMT (k=16).^30, 33, 35, 47, 50, 51, 56–58, 67, 69, 72, 80, 84, 91, 112^ ^30, 33, 35, 47, 50, 51, 56–58, 67, 69, 72, 74, 80, 84, 91, 112^. qMT parameters included the R1free (k=7)^30, 33, 35, 47, 50, 56, 69^ or T1free (k=5)^51, 58, 67, 80, 91^ including under saturation (T1sat, k=2),^57, 58^ T2free (k=4)^33, 35, 47, 69^ and T2bound (k=5),^33, 35, 47, 51, 69^ kf (k=8)^30, 33, 56, 67, 69, 72, 80, 91^ including under saturation (ksat, k=2),^57, 58^ the equilibrium magnetisation of the “bound” pool and the non-ideal inversion of the “free” pool signal (M0f and Sf, respectively, k=2),^30, 56^ f (k=3),35,47,51 and F (k=2).^30, 33, 47, 50, 56, 69, 72, 112^

#### 3.4.2. MT values across the brain

Studies varied as to the brain tissues in which MT was evaluated (Figure 1D and Table 2). Metrics were most often investigated in white matter (k=55)^7, 30–32, 35, 39– 47, 49, 51, 52, 54–58, 60, 62–64, 67–69, 71, 72, 74, 76, 77, 80–84, 89, 91–95, 97, 101–105, 107, 111, 112, 114^ and lesions (k=58),^7, 30–33, 35–37, 39–44, 46, 47, 49, 51, 56, 58, 60, 62, 67, 69, 74, 76–78, 80, 81, 83, 84, 86–96, 99–109, 111–114^ followed by grey matter (k=30),^7, 30, 32, 33, 35, 37, 38, 41–44, 48, 51, 52, 54, 56, 60, 61, 63, 64, 68, 71, 72, 74, 77, 89, 97, 105, 107, 114^ whole brain (k=19)^7, 34, 36, 46, 50, 60, 65, 75, 78, 85–88, 96, 98, 105, 107, 108, 110, 113, 114^ and specific regions of interest (k=22).^30, 31, 33, 35, 37, 41–43, 46, 49, 53, 56, 59, 64, 66, 70, 73, 74, 79, 83, 84, 89^ However, the definition of tissue categories varied. A distinction was often (but not always) made between ‘normal-appearing’ tissue and lesional tissue. Certain studies sub-divided tissue-type into lobes (e.g. frontal white matter) or regions of interest (e.g. deep versus cortical grey matter).

#### 3.4.3. MTR in RRMS and healthy controls

##### 3.4.3.1. Meta-analysis

Studies which compared MTR cross-sectionally between RRMS patients and controls (k=23 with available data) were submitted to a random-effects meta-analysis, with brain region as a nested factor. Irrespective of brain region, MTR for RRMS patients was on average 1.1 percent units [95% CI -1.47pu to -0.73pu] lower than controls (z-value: -6.04, p<0.001, Figure 2). Between-study heterogeneity was high (Total *I^2^*=47.7%).

**Figure 2:**
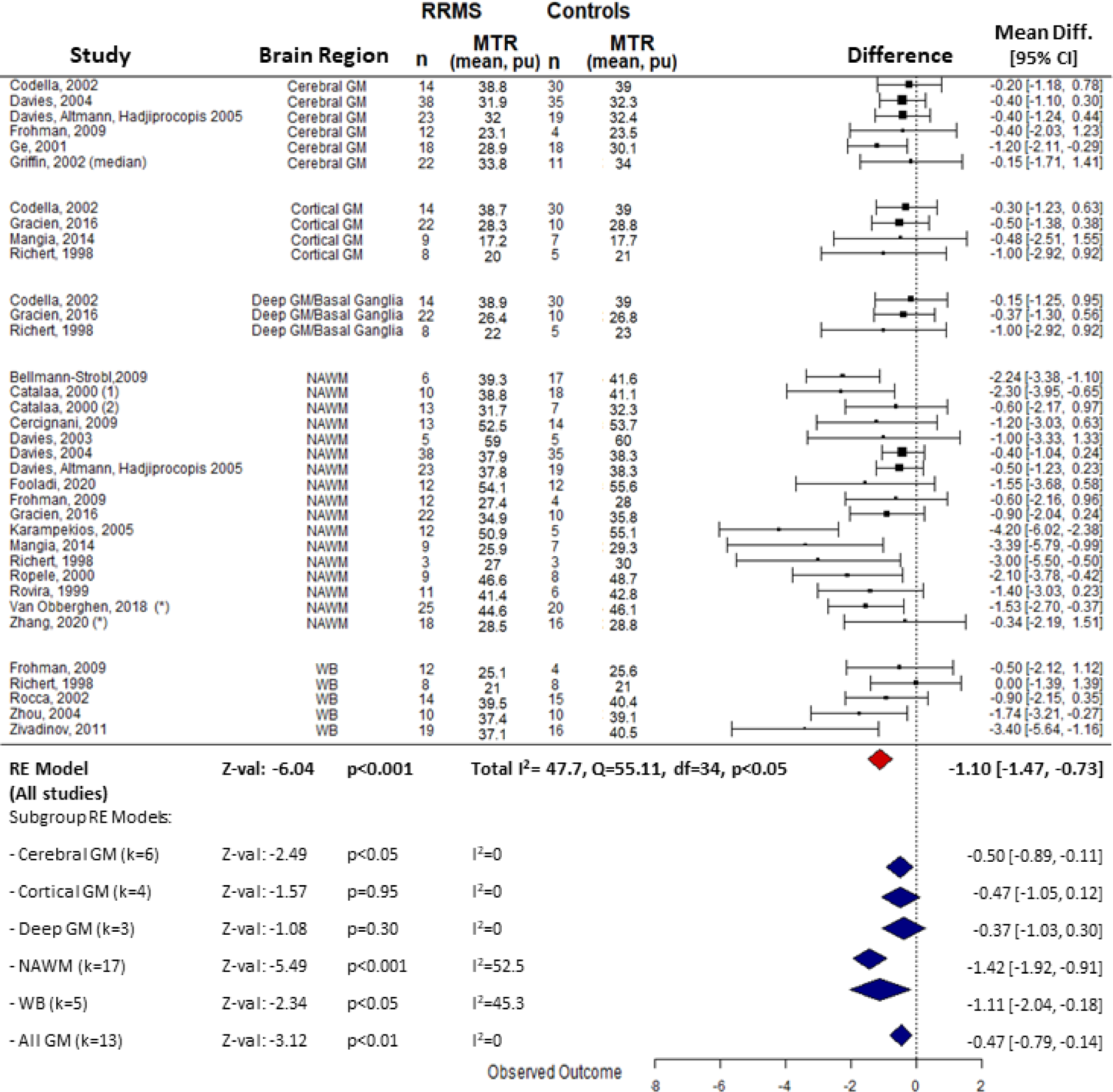
Random-effects meta-analysis of the difference in mean MTR between relapsing-remitting multiple sclerosis patients and control subjects with brain region as a nested factor. Study baseline data were used and sub-region estimates are shown below (uncorrected for multiple comparisons) including a grey matter model with sub-region of grey matter as a nested factor. GM: grey matter; NAWM: normal-appearing white matter; RE: random-effects; RRMS: relapsing-remitting multiple sclerosis; WB: whole brain;. *Averaged over sub-regions.

##### 3.4.3.2. Whole brain MTR

Whole brain MTR was measured in nineteen studies^34, 36, 46, 60, 65, 75, 78, 85– 88, 96, 98, 105, 107, 108, 110, 113, 114^ although eight did not distinguish between normal-appearing brain tissue (NABT) and lesions.^46, 65, 75, 85, 98, 105, 108, 110^ Average MTR in whole brain (k=9) was 35.58%^36, 60, 65, 78, 86, 87, 96, 107, 108^ with wide inter-study variance (range: 25.1%^60^ to 48.44%^36^, Figure 1E). Of the ten studies which made comparisons with healthy controls, sub-group meta-analysis showed that whole brain MTR was significantly lower for patients than controls with an absolute mean difference of - 1.11pu [95% CI -2.04pu to -0.18pu] (p<0.05, z-value 2.34, Figure 2 subgroup, k=5 with sufficient reported data) with moderate between-study heterogeneity (*I^2^*=45.3%).

##### 3.4.3.3. Normal-appearing white matter (NAWM) MTR

MTR of white matter (WM) was investigated in a large number of studies (k=48).^31, 32, 39–47, 49, 51, 52, 54, 55, 57, 58, 60, 62–64, 67, 68, 71, 74, 76, 77, 80–84, 89, 91–95, 97, 101, 103–107, 111, 114^ Typically, WM was defined as whole brain NAWM, with some exceptions such as regions of interest (ROIs) of NAWM contra-lateral to lesions of similar size,^32, 40, 91^ “dirty-appearing” WM^67, 94^ and NAWM sub-regions^51, 55, 64, 80, 82–84, 92, 93, 106^ (e.g. lobar WM,^41, 42, 103, 104^ NAWM close to cortical grey matter,^46^ perilesional NAWM^49, 91, 95^).

The mean NAWM MTR across studies was 69% (k=32)^31, 32, 39, 40, 45– 47, 51, 52, 54, 58, 60, 63, 67, 71, 77, 80, 81, 83, 84, 89, 92–95, 97, 101, 104–107, 111^ (range: 25.95%71 to 84%,104 Figure1E).

Overall, NAWM MTR was lower in RRMS patients compared with healthy _controls,_39,45,46,52,53,55,58,62-64,67,71,76,77,80,83 _although some studies found no_ difference^41, 43, 44, 47, 51, 60, 81, 84^. One study reported lower MTR in controls than patients.^74^ Random-effects sub-group meta-analysis (Figure 2) showed MTR of NAWM in RRMS was significantly lower than controls, with an absolute mean difference of -1.42pu [95% CI -1.92 to -0.91] (z-value -5.49, p<0.001, k=16 with sufficient data^v^) and considerable between-study heterogeneity (*I^2^*=52.5%).

##### 3.4.3.4. Grey Matter MTR

Twenty-three studies investigated grey matter MTR.^32, 37, 38, 41, 43, 44, 48, 51, 52, 54, 60, 61, 63, 64, 68, 71, 74, 77, 89, 97, 105, 107, 114^ Mean cerebral normal-appearing grey matter (NAGM) MTR was 31.5% (k=9),^48, 52, 54, 60, 61, 64, 97, 105, 107^ and consistently lower NAWM MTR^54, 64, 105^ with a wide range (Figure 1E). Cortical NAGM MTR, for example, was 2.9pu lower when using a bSSFP sequence compared with a GRE sequence within the same cohort.^89^

A random-effects sub-group meta-analysis (Figure 2) showed MTR to be significantly lower for RRMS patients than healthy controls across all grey matter regions (absolute mean difference -0.47 [95% CI -0.79 to -0.14], z=-3.12, p<0.01, k=13) with low between-study heterogeneity (*I^2^*=0%). When grey matter sub-regions where separately probed, follow-up random-effects models showed a significant difference for cerebral (mean difference -0.50, z-value -2.49, p<0.05) but not cortical or deep grey matter (mean difference -0.47 and -0.37, z-value -1.57 and -1.08, p=0.95 and 0.30, respectively, Figure 2). However, other studies (which did not report effect sizes) did not find between-group differences in MTR within cerebral^44, 51^ or cortical NAGM,^41, 43^ or within the basal ganglia.^41, 43^ Moreover, sub-regional variation was reported. For example, grey matter MTR in the parieto-occipital lobes, but not other regions, was lower for patients than controls in one study,^64^ and voxelwise differences in the left posterior cingulate cortex, right orbitofrontal cortex, bilateral insula and lenticular nuclei were noted elsewhere between patients and controls.^38^

##### 3.4.3.5. Lesion MTR

Forty-nine studies measured MTR in lesions.^31, 32, 36, 37, 39–44, 46, 49, 51, 58, 60, 62, 67, 74, 76– 78, 80, 81, 83, 84, 86–90, 92–96, 99–101, 103–109, 111–114^ MTR was nearly always lower in WM lesions than in NAWM (k=23, Figure 1E),^31, 40, 43, 46, 47, 51, 58, 62, 67, 71, 74, 77, 81, 83, 89, 91, 94, 95, 101, 103, 104, 106, 111^ “dirty-appearing” WM94 and healthy control WM (k=4).^39, 43, 81, 84^ Cortical lesion MTR was also lower than cortical NAGM.^89^ However, there was some regional heterogeneity. WM lesion MTR (and ihMTR) was not significantly lower than NAWM in the corpus callosum^83^ nor when several NAWM ROIs were combined.^84^

There was clear variation in MTR across lesions (Figure 1E), partially dependent on lesion characteristics,^43, 90^ which varied across the literature. In particular, MTR in T1- weighted ‘black holes’ was lower than in T1-weighted-isointense, T2-weighted visible lesions^104, 105^ although not always significantly.^106^ There was not typically a significant difference between MTR in contrast-enhancing lesions (CELs) such as nodular- enhancing CELs, and non-CELs,^90^ ‘pure T2-w lesions’ or T1 ‘black holes’.^104^ However, ring-enhancing CELs showed lower MTR than densely-enhancing^80^ or nodular-enhancing CELs.^81^ In addition, interdependency between lesion volume and MTR was reported,^43, 46^ although results are mixed.^96^

##### 3.4.3.6. MTR in other sub-regions

Seventeen studies measured MTR in other sub-regions of the brain (n=17)^31, 37, 41– 43, 46, 49, 53, 59, 64, 70, 73, 74, 79, 83, 84, 89^ including the thalami,^37, 41, 43, 53, 64, 73, 83, 84, 89^ putamen,^37, 41, 43, 64, 83, 89^ caudate nuclei,^37, 41, 43, 64, 89^ corpus callosum,^64, 70, 83, 84^ internal capsule,^46, 64, 83, 84^ globus pallidus,^37, 41, 43, 89^ cerebellum,^42, 79^ hippocampi,^73, 89^ cerebral corticospinal tract,^59^ accumbens,^89^ amygdala,^89^ cingulate cortex,^73^ and parietal cortex.^73^

A random-effects meta-analysis with brain sub-region as a nested factor showed no significant difference in baseline MTR between patients and controls (absolute mean difference -3.31pu [95% CI -8.65, 2.03], z-value=-1.23, p=0.215, Figure 3). Although between-study variance was low (*I*^2^=0.07%), total model variance was high (*I*^2^=98.9%) due to high variation in brain region (Figure 1E).

**Figure 3:**
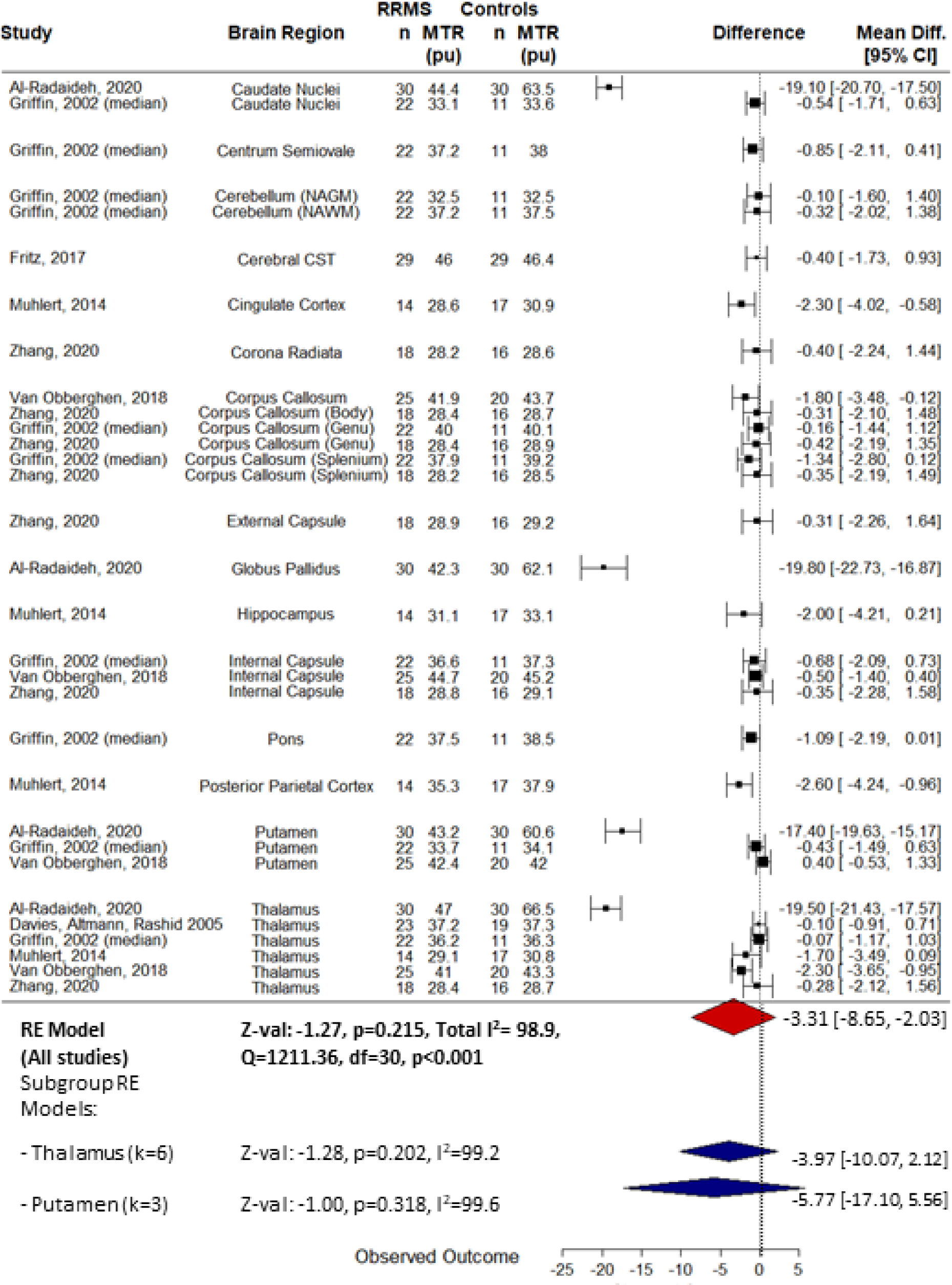
Random-effects meta-analysis to assess the difference in MTR in sub-regions across the brain between patients with relapsing-remitting multiple sclerosis and healthy controls. Study baseline data were used. CST: corticospinal tract; RE: random-effects; mean diff.: absolute mean difference in MTR; 95% CI: 95% confidence interval.

Since the number of studies examining MTR for most individual brain regions was low (*k*<3), follow-up sub-group random-effects meta-analyses were only performed for the thalamus (k=6) and putamen (k=3). There was no significant difference in baseline thalamic MTR between RRMS patients and healthy controls (mean difference -3.97pu [95% CI -10.07 to 2.12], z-value=-1.28, p=0.202, Figure 3) and high between-study variance (*I*^2^=99.2%). One additional study also found no difference in thalamic MTR between patients and controls (no effect size reported).^41^ Similarly, for the putamen, there was no difference between patients and controls (mean difference -5.77pu [-17.10 to 5.56], z-value=-1.0, p=0.318]) and heterogeneity was high (*I*^2^=99.6%). High between-study heterogeneity may be explained by differences in MT sequences used.^89^

#### 3.4.4. Longitudinal MTR change and therapeutic response

Thirteen studies assessed longitudinal change in mean MTR in one or more brain regions, with a maximum of three years follow-up. A linear mixed-model revealed that time did not have a significant effect on MTR when all brain regions were considered (β=-0.14 [-0.9 to 0.61], t-value=-0.38, p=0.71, Supplementary Table 5 and Supplementary Figure 1).

##### 3.4.4.1. Longitudinal change in whole brain MTR

Ten studies examined the longitudinal evolution of whole brain MTR^34, 36, 87, 88, 96, 98, 107, 108, 110, 113, 114^ of which five reported sufficient data to estimate longitudinal change in NABT MTR.^36, 87, 96, 107, 108^ A linear mixed-model showed that time did not significantly predict NABT MTR (β=-0.117 [-0.21 to -0.02], t-value=-2.65, p=0.019, Supplementary Table 6).

Nevertheless, individual studies reported small (e.g. <1% absolute change over two years^47^) but significant longitudinal decline in whole brain MTR.^36, 98^ A slower (non-significant) MTR decline (e.g. ∼0.02% every two months over 14 months^96^) and inter-subject variation were also reported. ^88, 98^ Additionally, longitudinal stagnation or increase in MTR with treatment compared with longitudinal decreases in MTR in placebo arms was evident in large, placebo-controlled cohorts over two years,^108, 114^ suggesting MTR as a putative therapeutic endpoint. However, one study reported no deterioration in whole brain MTR with glatiramer acetate treatment but lacked validation against a placebo-arm.^87^

##### 3.4.4.2. Longitudinal change in NAWM MTR

Fifteen studies examined the longitudinal evolution of NAWM MTR^39, 40, 43–45, 54, 62, 81, 91– 93, 101, 107, 111, 114^ of which seven reported appropriate data for a linear mixed-model to assess longitudinal change; NAWM did not change significantly over time (β=-0.082 [-0.13 to -0.29], t-value=0.78, p=0.44, Supplementary Table 7).^39, 40, 81, 91–93, 107^

In studies which reported a significant change over time, and in line with a previous report,^111^ absolute change in NAWM MTR was small (<1.5% up to 36 months) with reported estimates of an annual decline of 0.1% in early RRMS, possibly preceding clinical onset by years.^54^ However, others found no change in NAWM MTR over two years in an early MS cohort with minimal disability, after controlling for age and gender.^43^ Alternatives to the arithmetic mean such as histogram peak location may, nevertheless, reveal changes over 12-32 months.^45^

##### 3.4.4.3. Longitudinal change in grey matter MTR

A linear mixed-model of all brain regions suggests no effect of time on NAGM MTR but there were insufficient data for follow-up analyses (see section 3.4.4). In literature, however, MTR in grey matter decreases gradually (∼0.18pu annually, compared with 0.01pu in controls),^54^ although perhaps faster than NAWM MTR in RRMS.^54^ However, over two years, such gradual decline is not statistically significant.^43^ The longitudinal rate of grey matter change is unaffected by anti- phospholipid antibody (APLA) status,^107^ or treatment with IfN-β^54^ or laquinomod, ^114^ although the latter may slow decline initially.

##### 3.4.4.4. Longitudinal change in sub-regional MTR

There was no evidence of longitudinal change in MTR when all brain regions were considered (see section 3.4.4). Since there were few studies examining each brain sub-region (Supplementary Figure 1), no further meta-analyses of longitudinal change in MTR within brain sub-regions were constructed. However, no significant longitudinal change in MTR has been found in the thalamus, putamen, pallidum or caudate over two years.^43^ Separately, despite significant change in thalamic MTR (-

0.13pu/year) over two years, this was not significantly different from the rate of change in control thalamic MTR,^53^ and did not differ between those patients who were or were not treated with IfN-β

##### 3.4.4.5. Longitudinal change in lesion MTR

A linear mixed-model showed that lesion MTR did not change significantly longitudinally (β=0.375 [-0.56 to 1.30], t-value=0.82, p=0.42, Supplementary Table 8).^36, 40, 81, 87, 91–93, 96, 107, 111^ However, MTR longitudinal evolution depends on lesion characteristics^43^ and may be subtle^88^ (Supplementary Figure 1 and 2). MTR of active CELs varies from month-to-month before and after enhancement,^62, 91–93, 101, 112^ while MTR of GM lesions,^43^ ‘slowly expanding’ lesions,^99^ T1-weighted hypointense,^87^ and T2-weighted hyperintense^87, 96^ lesions may remain relatively stable over several years, irrespective of relapses.^96^

Increases in lesion MTR may also occur,^81^ such as within non-expanding lesions, although this may be accompanied by changes in T1^99^ and/or lesion load^113^. MTR increases may be seen with treatment (e.g. fingolimod^40^ over two years) although not always (e.g. laquinomod^114^). Steroids can increase CEL MTR^93, 101^ although certain DMTs, including delayed-release dimethyl fumarate^108^ or IfN β-1b^101, 109^ do not appear to alter CEL MTR. Furthermore, CELs do not tend to recover to NAWM MTR values^31, 93, 111^, and their longitudinal evolution may be predicted by the change in MTR of the first month post-enhancement.^93^ MTR in reactivated CELs also may deviate from NAWM MTR to a greater extent than new CELs.^91^

MTR fluctuations in lesions has been partially ascribed to low reproducibility, changes in interstitial water due to acute inflammation, or perhaps remyelination.^32^ Yet, when mixed lesion types are considered, a longitudinal global MTR decrease is typical.^43, 44^

#### 3.4.1. Clinical correlates of MTR

Eight studies reported correlation coefficients between MTR and EDSS permitting a meta-analysis (with brain region as a nested factor) to be performed. There was a significant negative association between EDSS and MTR across all brain regions; *r*=-0.30 [95% CI -0.48 to -0.08] (z-value=-2.91, p=0.01, Figure 4) and between-study heterogeneity was low (Total *I^2^*=0%). Across individual studies, sub-regional results were mixed but in general, suggest that there is no association between EDSS and _MTR._83,89

**Figure 4:**
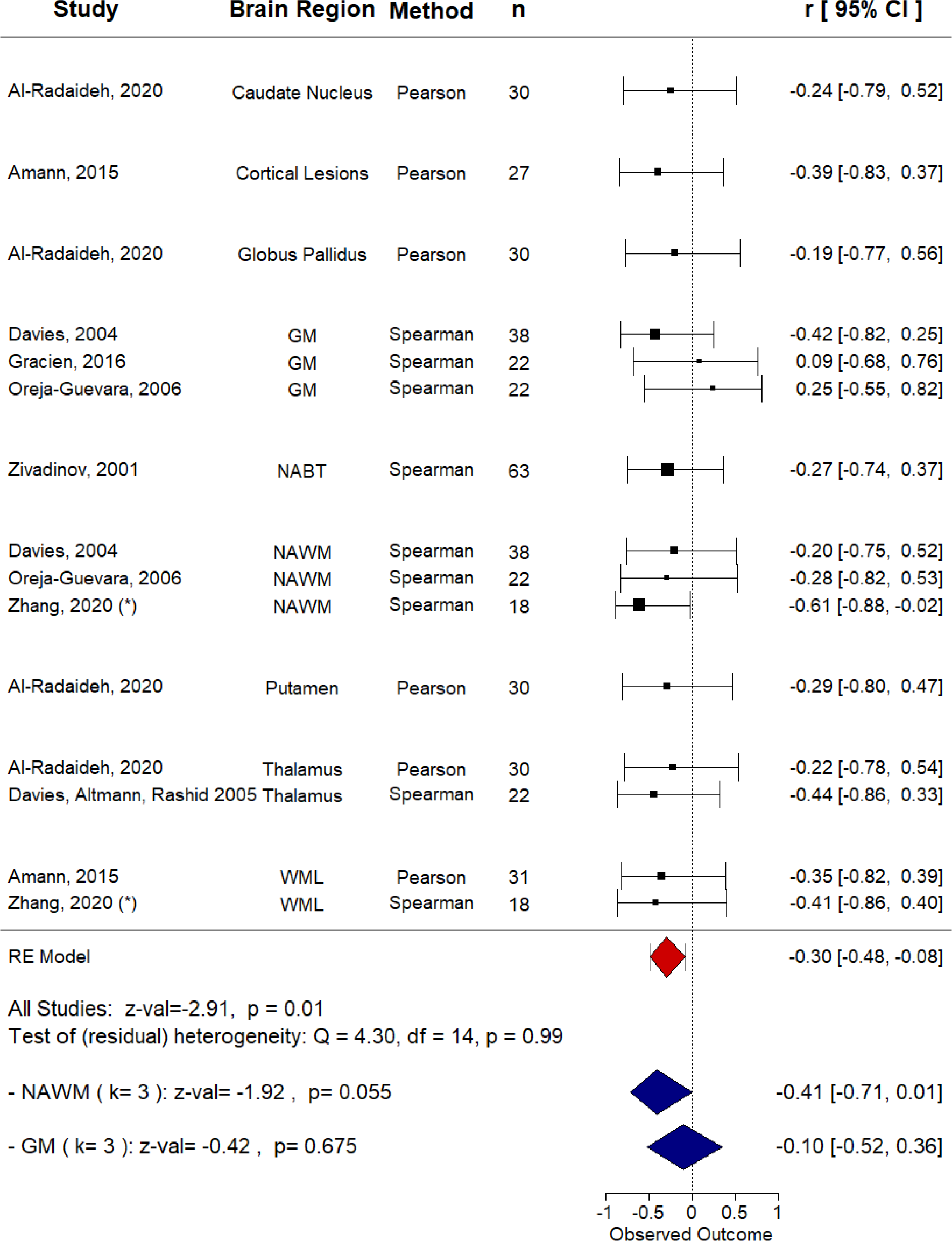
Meta-analysis of association between MTR and clinical disability in relapsing-remitting multiple sclerosis. Clinical disability was defined as Expanded Disability Status Scale (EDSS) score. Studies (k=8) were submitted to a multi-level random-effects model with brain region as a nested factor within each study. For brain regions in which k>2, random-effects sub-group analyses were also performed. * MTR values were averaged over sub-regions of NAWM. Studies which did not report a correlation coefficient were not included. GM: grey matter; NABT: normal-appearing brain tissue; NAWM: normal-appearing white matter; WML: white matter lesions; RE: random effects; CI: confidence interval.

##### 3.4.1.1. Whole brain MTR and clinical correlates

In terms of whole brain MTR clinical correlates, there is some evidence that NABT MTR correlates with EDSS^86^ (Figure 4) but not retinal nerve fibre layer (RNFL) thickness or low letter contrast acuity.^60^ NABT MTR may predict longitudinal memory decline and, in combination with brain parenchymal fraction and two year change in ventricular fraction, information processing speed over seven years.^36^ No such association was found between NABT MTR and verbal fluency.^36^ However, this study was limited by the lack of comparative longitudinal control data. Furthermore, longitudinal evolution of NABT MTR does not appear to depend on APLA status of patients.^107^

##### 3.4.1.2. NAWM MTR and clinical correlates

Many studies examined the relationship between clinical disability and NAWM MTR (Table 2), yet only three studies reported effect sizes. A sub-group meta-analysis for NAWM showed a borderline negative association between EDSS and NAWM MTR (p=0.055, *r*=-0.41 [95% CI -0.71 to -0.01], Figure 4) with low between-study variance (*I^2^*=0%). However, the small number of studies (k=3) limits the generalisability of this finding, particularly given under-reporting of non-significant effect sizes. Indeed, all ten studies which examined the association between NAWM MTR and EDSS found no association,^39, 45, 52, 54, 71, 84, 87, 89, 103^ although one study reported a significant correlation between baseline NAWM MTR and change in EDSS over eighteen months (but not baseline EDSS).^97^

Evidence of relationships between NAWM MTR and other clinical measures was mixed. For example, NAWM MTR was associated with MSFC z-score at 24 month follow-up but not baseline^39^ while, separately, there was no relationship between MSFC z-scores and NAWM MTR^71^ or two year change in NAWM MTR.^54^ Associations may also be region- and model-dependent; for example, temporal lobe MTR was one of several significant predictors of MSFC and SDMT (an attention test) scores, independently, in regression models.^41^

In terms of other biomarker correlates, WM MTR was weakly associated with serum neurofilament – a marker of neuronal injury - in RRMS (although not in control subjects), adding to evidence validating MT imaging as a biomarker of myelin integrity.^68^ NAWM MTR does not however appear to be related to RNFL thickness or low contrast letter acuity.^60^

#### 3.4.1.3. Grey matter MTR and disability

Eight studies examined the relationship between grey matter MTR and EDSS (Table 2) with some demonstrating significant associations^52, 61^ and others finding no such relationship.^38, 54, 63, 71, 89^ One study found an association between baseline grey matter MTR and change in EDSS, but not baseline EDSS.^97^ A follow-up sub-group random-effects meta-analysis showed no significant association between study baseline grey matter^vi^ MTR and EDSS (p=0.675, *r*=-0.10, [95% CI -0.52 to 0.36], Figure 4) and low between-study heterogeneity (*I^2^*=0%), but the number of studies was small (k=3).

Four studies examined the relationship between grey matter MTR and the MSFC.^38, 52, 54, 71^ MSFC z-score did not correlate with cerebral NAGM,^52^ cortical NAGM^71^ or voxels of NAGM for which the MTR differed from controls.^38^ Furthermore, neither change in MSFC nor its cognitive component correlated with change in MTR in NAGM over two years.^54^

Regarding other clinical variables, NAGM MTR was significantly correlated with age^89^ as well as RNFL thickness of eyes affected by optic neuritis.^60^ Female subjects may also have higher NAGM MTR^52^ although this was not a consistent finding.^89^ In addition, NAGM MTR correlates with T1 and myelin water fraction.^74^ On the other hand, grey matter MTR did not correlate with low contrast letter acuity,^60^ RNFL of eyes unaffected by optic neuritis,^60^ serum neurofilament levels,^68^ immune cell BDNF secretion,^105^ APLA status,^107^ fatigue,^48^ or disease duration.^38, 52, 89^ Change in NAGM MTR was not associated with relapse rate, baseline T2 lesion volume or change in T2 lesion volume over two years^54^ nor APLA status over three years.^107^

##### 3.4.1.4. MTR in other sub-regions and disability

MTR within other sub-regions such as the internal capsule,^46, 83^ cerebral corticospinal tract,^59^ caudate, pallidum, putamen, accumbens, hippocampus and amygdala,^89^ and corpus callosum^83^ was not associated with EDSS. There was a negative association between thalamic MTR and EDSS averaged over two years,^53^ although two year change in thalamic MTR was not associated with EDSS at follow-up,^53^ possibly reflecting a lack of change in thalamic MTR over two years.^43^

Regarding other clinical correlates, no relationship was found between thalamic MTR or rate of change of MTR over two years and MSFC.^53^ Nevertheless, the walk component of the MSFC was negatively associated with thalamic MTR.^53^ In the cerebral corticospinal tract, MTR was associated with walk velocity and Two Minute Walk Test but not Pyramidal Functional Systems Score, gender or symptom duration, but perhaps slightly dependent on age.^59^ MTR of the corpus callosum was positively associated with PASAT (the cognitive component of the MSFC) score, although possibly mediated by lesion load.^70^ Cognitively impaired RRMS patients may also have marginally reduced MTR in the corpus callosum compared with unimpaired patients.^70^ There may be an influence of age on MTR in the basal ganglia, thalamus and hippocampus.^89^ Finally, MTR in an area of the cerebellum thought to be involved in movement trajectories was associated with performance on the MSFC arm component.^79^

##### 3.4.1.5. Lesion MTR and clinical correlates

In lesions, any relationship between clinical disability and MTR is at most weak.^37, 39, 41, 49, 84, 89, 103^ Only two studies reported a correlation coefficient (Figure 4) for an association with EDSS and hence a meta-analysis was not performed for lesion MTR alone.

This relationship may depend on lesion type, characteristics^42^ and location.^89^ For example, cortical, but not white matter, lesion MTR was related to EDSS, after adjusting for demographic factors.^89^ Furthermore, when lesions were grouped according to their inflammatory and neurodegenerative characteristics, lesions with low MTR were found to predict attention deficits (SDMT) and general disability (MSFC), when combined with age and depression score.^42^

The timescale of the study, disease duration^89^ and treatment of confounding variables may affect the strength of association. A longitudinal relationship between MTR in lesions and clinical disability developed with longer disease duration in one study when not present at baseline ^39^. Lesion MTR, when combined with T2- weighted lesion and NAWM measures, was also related to longitudinal change in deambulation (MSFC T25FW).^43^ However, baseline T2-weighted lesion MTR was not a significant predictor of change in memory, verbal fluency or information processing speed over seven years.^36^

More generally, the association between MTR and clinical disability may depend on which clinical measure(s) are used. For example, lesion MTR was not significantly different between cognitively impaired and unimpaired patients, when assessed by an extensive battery of neuropsychological tests.^86^ Similarly, MTR within (mixed-type) lesions did not correlate with motor tasks (finger tapping rate or 9HPT),^78^ and was not a significant predictor in regression models to predict general clinical disability (MSFC), attention (SDMT) or fatigue (Fatigue Scale for Motor and Cognitive functions).^41^

Some studies indicate associations between MTR as a measure of myelin integrity and other imaging markers of disease in MS. Weak evidence suggests that the uptake of radiotracer ^18^F-PBR111, which binds to the 18-kD translocator protein, is greater in around 60% of T2-w FLAIR hyperintense regions compared with non-lesional regions with high MTR.^49^ Higher uptake of ^18^F-PBR111 is suggestive of a pathological increase in macrophages and microglia. Single-subject MR spectroscopy has shown elevated choline and lactate/lipids suggestive of demyelination and injury to cell membranes, alongside decreases in N-acetyl compounds, creatine and myoinositol indicating axonal loss and increased glial cell infiltration, compared with NAWM in a tumefactive CEL.^31^ MTR in lesions is strongly associated with other imaging metrics such as MMC,^112^ and kf ^67, 80, 112^ and, to a lesser extent, quantitative T1^67, 74, 112^ and myelin water fraction.^74^ Lesion MTR is negatively correlated with relative activation on functional MRI in motor areas suggestive of functional adaptations to loss of myelin integrity, although perhaps confounded by lesion volume.^78^ MTR correlates weakly with diffusion-weighted imaging metrics including fractional anisotropy^95^ in large T2-w lesions and mean diffusivity^103^ in chronic lesions, but not significantly with susceptibility-weighted phase imaging values, despite a negative trend.^103^ Additionally, T2-w and T1-w ‘black hole’ lesion volume, as well as two year change in T2-w lesion volume may predict lesion MTR thirteen years later, although uncorrected for baseline lesion MTR.^113^

Nevertheless, as a general trend across the RRMS literature, MTR within lesions does not tend to correlate with other disease biomarkers. T2-w lesion MTR is not significantly associated with age,^89, 103^ time since diagnosis,^37^ visual contrast acuity or RNFL thickness,^60^ immune cell BDNF (brain-derived neurotrophic factor) secretion,^105^ or APLA status (+/-).^107^ MTR in CELs was not associated with anti-CD3 plus anti-CD28 stimulated BDNF secretion, despite a negative trend.^105^ MTR in T1-w ‘black holes’ is not associated with RNFL thickness or visual contrast acuity.^60^ There is some evidence that APLA+ patients show greater reduction in MTR in T1 ‘black holes’ compared to APLA- patients over three years, but this may be driven by lesion volume changes.^107^ Evidence for associations between lesion MTR and disease duration or gender is mixed, and may depend upon acquisition parameters and lesion type.^89, 103^

#### 3.4.2. MTsat

Three studies used MTsat (Figure 1C),^7, 66, 102^ beginning with Helms et al. ^7^ who showed that, on a whole brain histogram, the WM MTsat mode appeared visually reduced in a RRMS patient compared with controls. Furthermore, compared to NAWM, MTsat in a CEL and non-enhancing lesions was visually lower on a parametric map.^7^

Saccenti et al.^102^ confirmed that MTsat was significantly lower in WM “plaques” and periplaques than NAWM. Yet, MTsat did not correlate with EDSS or disease duration in plaque, periplaque or NAWM ROIs.^102^ MTsat may additionally correlate with radial diffusivity, T1w/T2w ratio and synthetic MR-derived myelin volume fraction, although this was stronger in plaques than NAWM.^102^

Finally, Kamagata et al.^66^ used MTsat as a surrogate for myelin volume fraction to calculate the tract-averaged MR g-ratio within WM in a small RRMS cohort.^66^ The g-ratio was increased (indicating myelin degradation and/or axonal loss) compared with healthy controls, in motor somatosensory, visual and limbic regions. Subnetwork g-ratio strongly negatively correlated with WM lesion volume, but not with disease duration or EDSS, although the latter was correlated with g-ratio connectome nodal strength mainly in motor, visual and limbic regions.

#### 3.4.3. ihMTR

Two studies employed ihMTR as a measure of myelin status in RRMS.^83, 84^ ihMTR was reduced in lesions and NAWM compared to control WM, and reduced in lesions compared to NAWM.^84^ Within sub-regions, single-slice ihMTR was lower for patients in the thalamus, frontal, temporal and occipital lobes compared with controls, but no different in the corpus callosum, internal capsule or putamen.^83^ ihMTR varied across WM tracts, but was highest in the internal and external capsule and lowest in the genu of the corpus callosum.^83, 84^ ihMTR in WM lesions, but not NAWM, was negatively associated with EDSS.^84^ However, when sub-regions were considered, EDSS was significantly associated with ihMTR (but not MTR) in frontal and temporal NAWM, the corpus callosum, internal capsule and the thalami.^83^

#### 3.4.4. qMT

qMT metrics varied across studies (see section 3.4.1). Sled and Pike^33^ first modelled the compartmental MT signal in RRMS in two lesions on a single-slice PD-weighted image for a RRMS patient. Compared to frontal WM, lesions had reduced kf, F, R1free, and T2bound and increased T2free. Parameter estimates were higher for the newer lesion compared to the older lesion for kf, F and R1free, but lower for T2free and T2bound. Indeed, other studies also show lower kf and ksat lesions than NAWM and healthy control WM, while T1free and T1sat present the inverse pattern.^58, 67, 80^ Up to four months prior to the appearance of new or reactivating CELs, kf may even decrease while T1free increases.^91^ However, changes are subtle, and month-by-month change may be less predictable for reactivating CELs.

Increasing lesion severity coincides with decreasing kf^33, 67, 80, 91^ and ksat,^58^ while conversely T1fre ^67, 80^ and T1sat^58^ are elevated in acute, compared to mild, lesions.

However, dense CELs have higher kf but lower T1free values than ring CELs.^80^ F^30^, *f*^35, 51^, R1free,^30, 47^ and T2bound,^47, 51^ are also reduced in lesions compared to NAWM and control WM, with reduced F and R1free in T2 hyperintense lesions visible on SIR-derived parametric maps.^50, 56^. Finally, MMC is reduced in CELs but may recover post-enhancement.^112^ The relationship between pathology and qMT-derived metrics is evidently complex, but may still differentiate between lesions with similar MTR, particularly when lesions are T1-weighted isointense.^67^

Differences between NAWM and control WM qMT are, however, subtle. Some studies report differences for qihMT,^84^ T1free,^67^ F^47^ and kf ,^47, 67, 80^ while others show no differences for k ^33, 72^ F,^72^ f,^51^ T2bound,^51^ T1free,^80^ R1free,^47^ or qMT.^84^ Seven studies were thus submitted to a random-effects meta-analysis to compare qMT in NAWM and WM^33, 35, 47, 51, 58, 67, 80^ There was no significant difference between patients and controls across all qMT metrics (standardised mean difference -0.22 [95% CI -0.62 to 0.18], z-value: -1.12, p=0.25, Figure 5). Additional follow-up models for metrics where k ≥ 3 also showed no significant difference for T1free, T2bound and kf (α=0.0125, Figure 5) despite a trend for kf. Other brain regions were not assessed due to limited data.

**Figure 5:**
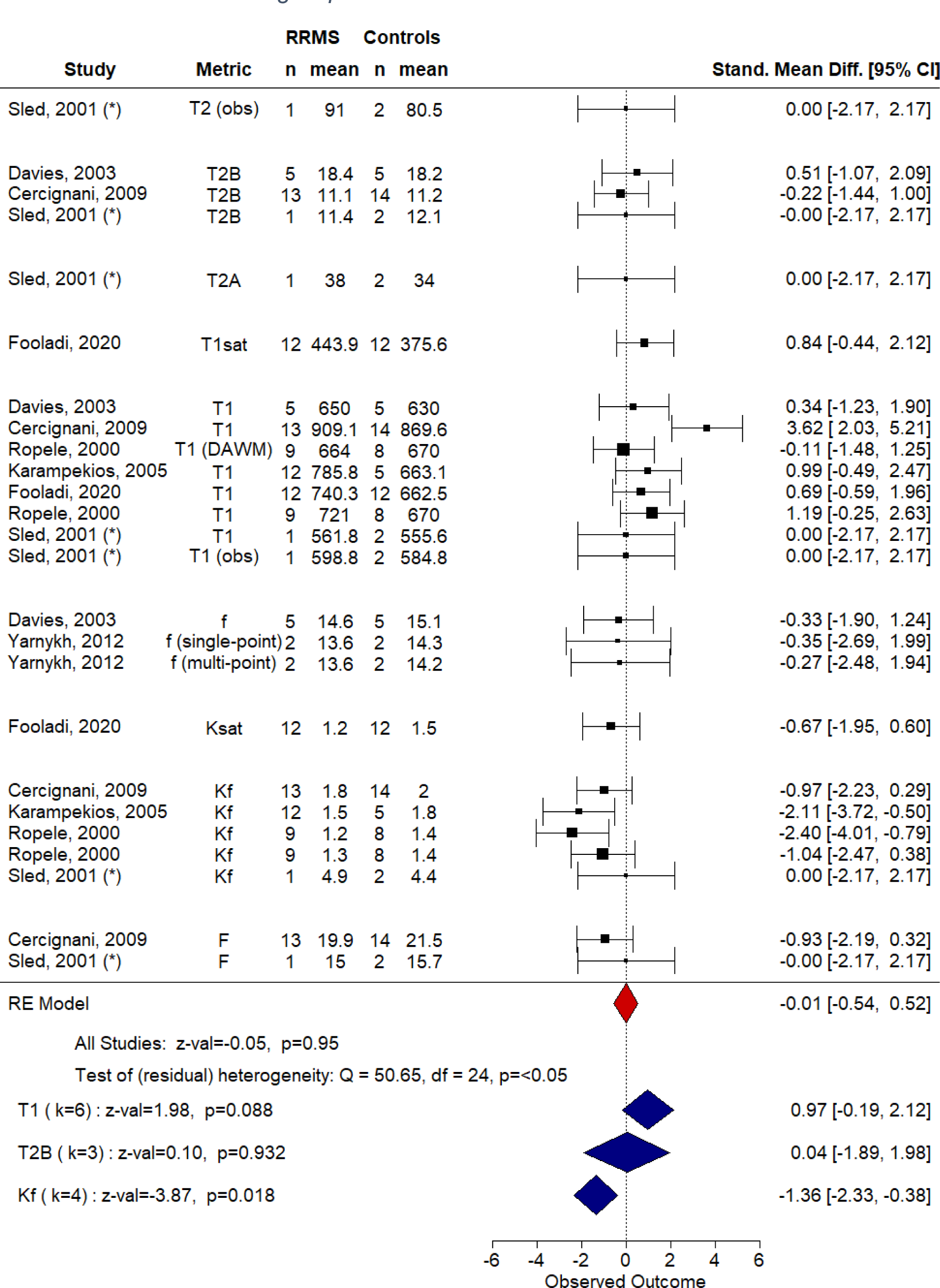
Random effects meta-analysis of magnetisation transfer compartmental model parameters in white matter. Metric was a nested factor within study and subgroup (e.g. DAWM versus NAWM) was nested within metric.R1 was converted to T1 for comparability where necessary. DAWM: dirty-appearing white matter; NAWM: normal-appearing white matter; Stand Mean Diff.: standardised mean difference. (*) frontal white matter; α=0.05 for omnibus test and α=0.05/4=0.0125 for sub-groups.

In cortical grey matter, kf, F, R1free and T2bound appear lower and T2free higher than in lesions and frontal WM.^33^ RRMS patients have lower kf than controls in cortical grey matter but F does not differ, except for patients with high disability.^72^ No differences between patients and controls were found in cerebral or cerebellar grey matter for *f*, T1free or T2bound.^51^ In deep grey matter, *f* was lower for patients than controls.^35^ However, differences in methodology can results in over- or underestimation of *f* in certain ROIs (e.g. thalami).^35^

Few studies have examined the relationship between qMT and clinical disability in RRMS. Cortical grey matter kf may be negatively associated with EDSS and Choice Reaction Time, but not SDMT or PASAT.^72^ Associations between EDSS and both qMT and qihMT in lesions, but not NAWM have also been reported.^84^ Combining qMT parameters, and including covariates such as lesion load and age may improve models^47^ but collinearity (e.g. between *f* and T2bound or kf and T1free) may be problematic if used in the same model.^51, 67^

## 4. Discussion

Our systematic review revealed 86 studies which used MTI to investigate cerebral RRMS pathology. The vast majority (87%) of these used MTR.

### 4.1. Common findings

Lesion MT was found to be lower than in NAWM. MT was also generally reduced in non-lesional brain for patients compared with healthy controls, indicative of subtle loss in microstructural integrity. Conversely, smaller sub-regions (e.g. thalamus, putamen) did not show such differences. Annual longitudinal decline in MT across all brain regions was subtle but inclined to fluctuate in lesions.

Although associations between MT measures and clinical disability in RRMS were apparent, relationships were weak, and confounded by factors such as age. This association may be limited by the lack of longitudinal data over sufficient time periods for divergence in disability to become apparent.

Studies examining longitudinal change and clinical correlates were limited to MTR; we did not identify any such studies using other techniques, such as MTR, ihMT or qMT.

### 4.2. Sources of bias

Analysis of the literature revealed: (1) the diversity of quantitative MTI acquisition protocols; (2) an affinity for MTR over alternative metrics; (3) general consistency in the directionality of MT changes in RRMS, but small effect sizes; and, (4) the challenge of clinical interpretation and comparison due to heterogeneous imaging characteristics.

#### 4.2.1. Sample Characteristics

Our search demonstrated a broad literature of MS-specific MTI studies, a considerable number of which were excluded due to lack of distinctions between MS sub-types, or grouped sub-types in analyses and results.

Overall, patient sample sizes across the RRMS MTI literature were small. Research with a technical or proof-of-concept focus tended to include a single subject or handful of participants (e.g. ^7, 30, 33, 35, 56, 106^). International clinical trials recruited larger cohorts (e.g. ^108, 110^) but at the expense of standardised, well-documented MTI protocols.

The absolute sensitivity of MT metrics to pathological changes in the brain of people with MS is modest; the difference in MTR between patients with RRMS and healthy controls is estimated to be small (∼0.5% to 2%) which is far lower than inter-study variability. Many studies were underpowered. Our review therefore highlights a need for validation of advanced, pragmatic quantitative MT techniques in larger cohorts, for reliable detection of disease effects.

Comparisons between MS and (typically) age-matched healthy control subjects featured in a number of studies, albeit often with significantly smaller control than patient groups. This is important as it can effectively provide reference data to improve comparability of MT metrics across studies and centres, provide an index of test-retest variance, and help to mitigate additional variability caused by technical differences in acquisition on different platforms. Control data may additionally help to account confounding variables associated with MTR, such as age^89^ and disease duration.^53^

Treatment effects are a potential confound of MT microstructure measures, and inter- and intra-study heterogeneity was apparent in DMT and steroid usage which is an additional source of variability. Although some studies control for treatment effects, greater consistency is required in studies whose primary focus is imaging biomarker validation.

Across studies, there was a near universal bias towards European and North American populations, which is likely to reflect geographical prevalence of MS, focus on the disease within healthcare systems, and access to MRI and research protocols.

Importantly, analysis of the location of study centres highlights possible bias due to data duplication from multiple or overlapping analyses of cohorts. This is rarely overtly reported, but may bias calculation of effect sizes.

#### 4.2.2. Imaging acquisition protocols

Systematic comparison of MTI in RRMS demonstrates substantial heterogeneity of MTI acquisition protocols. There was wide variation in magnetic field strength, pulse sequence, image weighting, excitation flip angle, TR and TE. With the rapid evolution of MRI hardware and techniques, such sources of variation are inevitable and well-recognised in the quantitative MRI literature. The nature of MT acquisition, however, makes MT measurements particularly sensitive to these factors. For example, simulations suggest that the difference between grey and WM MTR at 3T at an offset frequency of 1.5kHz is around 43% larger than at 1.5T.^82^ Use of proprietary hardware and pulse sequences allows broader access of MTI to research groups with limited MRI pulse programming expertise, but typically fixes, restricts and even conceals important pulse sequence parameters.

MT measurements are especially sensitive to characteristics of the MT pulse. Quantification typically assumes selective saturation of the ‘bound’ pool with minimal direct saturation of the ’free’ water pool. The extent to which this is achieved *in vivo* and the resulting tissue-type contrast, however, depends on the complex relationship between tissue properties, hardware, sequence parameters and MT pulse design features including the offset frequency, power, pulse duration and shape.^111^. In particular, our finding of wide variance in NAWM MTR in RRMS cohorts is suggestive of sequence parameter dependence. Early experiments with relatively low offsets (e.g. ^75, 95^) are likely to have a greater direct saturation effect. Improved harmonisation and standardisation of MT protocols between centres would help to minimise these sources of variability.

#### 4.2.3. Tissue types and definitions

Substantial variation observed in MTR values for different tissue-types is likely due not only to varying acquisition parameters discussed above, but also how tissue type is defined; and variations in methods by which the regions are segmented from structural imaging. For example, individual studies examine different combinations of WM, NAWM, cortical and deep grey matter structures, atlas-based regions of interest, and whole brain analyses. Moreover, MS “lesion types” in RRMS are defined by their signal characteristics; for example, T2-w hyperintensities, T1-w ‘black holes’, contrast enhancing lesions, and FLAIR hyperintensities.

#### 4.3. Implications for future studies using MT in RRMS

The findings of this review indicate potential for MT measures of microstructure as useful disease markers in MS, but also advocate for further validation against well-defined and meaningful clinical endpoints, and other biomarkers of MS disease activity and neurodegeneration in larger studies of adequate statistical power.

In order to achieve this, large highly characterised cohorts of patients with defined disease subtypes will need to be studied over time, using optimised MTI acquisition protocols to maximise sensitivity to pathological change. A pre-requisite is rigorous evaluation of technique reproducibility by test-retest measures in patients and healthy control subjects.

Sufficient cohort sizes for adequate statistical power to detect predicted effect sizes will inevitably require multicentre studies, and therefore optimisation and harmonisation of MTI protocols across multiple sites and MRI vendors and platforms, with assessment of inter-site variance and potential systematic differences in measures across centres. Adoption of more consistent definitions and methods for segmenting tissues of interest will also facilitate comparability across sites and studies. Improved MT methods may ultimately avoid the need for time-consuming segmentation, if whole brain analyses can provide useful disease biomarkers.

Examining change over several years is also key to assessing the trajectory of disease, and strategies for mitigating the effects of, for example, MRI system upgrades and changes in equipment during the course of longitudinal imaging studies need to be considered.

The majority of large-scale MT studies in RRMS to date have used MTR, which is relatively easy to acquire and analyse, but is highly sensitive to acquisition parameters, as well as T1 and B1 differences.

qMT provides the most accurate modelling of MT processes and is helpful for understanding MTI in healthy and pathological tissue, however the prolonged acquisition needed at multiple pulse powers and offset frequencies with adequate spatial resolution and whole-brain coverage is not currently feasible for clinical imaging in patients.

Emerging MT methods such as MTsat and ihMTR provide potentially more robust and specific measures of myelin integrity than MTR within clinically feasible acquisition times.^7, 115^ Histological validation in felines has shown that MTsat is sensitive to demyelination,^116^ and, in mice, ihMTR is more specific to myelin than MTR^115^ Both techniques, however, require further validation with histology and larger patient and healthy control cohorts.

We argue that, in order for MTI to evolve as a useful imaging tool in MS and other diseases, there is a need to establish consensus standards for image acquisition, analysis and reporting from an international group of experts working across centres, as has been successfully achieved with other quantitative MRI methods such as diffusion and perfusion imaging.^117–119^

### 4.4. Limitations and Conclusion

The scope of the present review is limited to studies of RRMS patients. Studies involving other MS subtypes were excluded, but may still provide relevant information on how MT metrics reflect disease activity. Similarly technical experiments in healthy volunteers crucial to the advancement of MTI were not included here. Meta-analyses did not further take into account patient or control group demographics. Finally and importantly, meta-analyses were limited by large inter-study heterogeneity and missing data.

In conclusion, this systematic review demonstrates the broad use of MTR in RRMS. The evidence evaluated suggests that MT imaging can detect subtle disease-related differences, however also highlights how large measurement variability due to differences in technique dominate over small effect sizes, which in turn limits clinical and biological interpretation. The implementation of more robust emerging quantitative techniques, and consensus regarding optimised, harmonised protocols in large well-characterised patient cohorts will be required to establish MTI as a useful microstructural marker in RRMS, for translation into wider clinical use.

## Supporting information

PRISMA 2020 Completed Checklist

## Data Availability

Extracted data may be provided upon reasonable request to the corresponding author(s).

## Acknowledgements

We are grateful to Dr Una Clancy for statistical graphics recommendations.

## Registration and protocol

This review was not registered and a protocol was not prepared.

## Availability of data

Extracted data may be provided upon reasonable request to the corresponding author.

## Competing interests

The authors declare no conflicts of interest relevant to this paper.

### Abbreviations

9HPT: nine-hole peg test
APLA: anti-phospholipid antibody
BDNF: brain-derived neurotrophic factor
bSSFP: balanced steady-state free precession sequence
CELs: contrast-enhancing lesions
CI: confidence interval
DMTs: disease-modifying drugs
EDSS: Expanded Disability Status Scale
FLAIR: fluid-attenuated inversion recovery
GRE: gradient echo
HASTE: Half-fourier Acquisition Single-shot Turbo spin-Echo
IfN-α/β: interferon-alpha/beta
ihMTR: inhomogeneous magnetisation transfer ratio
MS: multiple sclerosis
MTI: magnetisation transfer imaging
MSFC: Multiple Sclerosis Functional Composite
MTR: magnetisation transfer ratio
MTsat: magnetisation transfer saturation
NABT: normal-appearing brain tissue
NAGM: normal-appearing grey matter
NAWM: normal-appearing white matter
PASAT: Paced Auditory Serial Addition Test
PD-weighted: proton density-weighted
PPMS: primary progressive multiple sclerosis
qMT: quantitative magnetisation transfer
RNFL: retinal nerve fibre layer
ROI: region of interest
RRMS: relapsing-remitting multiple sclerosis
SAR: specific absorption rate
SDMT: Symbol-Digit Modalities Test
SE: spin echo
SIR: selective inversion recovery
SPMS: secondary progressive multiple sclerosis
T1-w: T1-weighted
T2-w: T2-weighted
T25FW: Timed 25-Foot Walk
TE: echo time
TR: repetition time
WM: white matter

## Footnotes

a Where only female subjects were recruited, the ratio was calculated as n/1. e.g. 15 F, 0 M would give F:M ratio of 15.

b Of patients whose data was analysed, where mean age was reported. If age was only reported for those recruited, this was still included. Median age was not included.

c Including Iran and Jordan

d The median of all reported cohort median EDSS at baseline or mean EDSS at baseline when median unreported

v One study (Catalaa, 2000) was included twice as separate protocols and cohorts were used, thus k=17

vi Note that, due to the small number of studies (k=3), cortical and cerebral grey matter were grouped together for this analysis.

