## Supplementary Material for "Quantitative magnetisation transfer imaging in relapsing-remitting multiple sclerosis: a systematic review and meta-analysis"

*Supplementary Table 1: Overview of studies which were excluded from the final selection due to mixed multiple sclerosis (MS) subtypes. CIS: clinically-isolated syndrome; MT: magnetisation transfer; MTR: MT ratio; MWF: myelin water fraction; PPMS: primary progressive MS; PSR: pool size ratio; qMT: quantitative MT; RRMS: relapsing-remitting MS; SPMS: secondary progressive MS.*

| Citation | Mixed Analyses | Subgroup Analyses | RRMS (n) | SPMS (n) | PPMS (n) | Other (n) | Healthy (n) | MT Metric |
|---|---|---|---|---|---|---|---|---|
| [1] | • | - | 16 | 1 | 1 | - | 9 | MTR |
| [2] | - | • | 13 | 10 | 15 | - | - | MTR; IR |
| [3] | - | • | 34 | 19 | - | 20 (CIS) | 16 | MTR |
| [4] | - | • | 11 | - | - | 17 (CIS) | 22 | MTR (-/+) |
| [5] | • | - | 59 | 12 | - | - | - | MTR |
| [6] | • | - | - | - | - | 18 (CIS) | 18 | MTR |
| [7] | • | - | - | - | - | 18 (CIS) | 18 | MTR |
| [8] | • | - | 11 | 3 | - | 4 (CIS) | 9 | PSR; $R1_f$ |
| [9] | - | • | 37 | - | 43 | - | 58 | MTR |
| [10] | - | • | 18 | 88 | - | - | - | MTR |
| [11] | • | - | 75 | | | | - | Fat sat. MTR |
| [12] | - | • | 56 (46) | 0 (8) | - | - | 56 | MTR |
| [13] | • | - | 77 | 8 | 3 | - | - | MTR |
| [14] | • | - | 41 | | | | 21 | MTR |
| [15] | - | • | 14 | 7 | - | - | - | MTR |
| [16] | • | - | 3 | 10 | | 5 (benign) | - | MTR |
| [17] | - | • | 22 | 6 | 4 | 1 (benign) | 27 | $f$; $T2_b$ |
| [18] | • | - | 18 | 32 | 25 | - | 129 | MTR |
| [19] | - | • | 50 | - | - | 50 (benign) | 10 | MTR |
| [20] | - | • | 20 | - | - | 19 (RIS) | 20 | MTR |
| [21] | - | • | 10 | 16 | 46 | 11 (benign) | 39 | MTR |
| [22] | - | • | 11 | 11 | 10 | 10 (benign) | 9 | MTR |
| [23] | - | • | 10 | 16 | 46 | 11 (benign) | 39 | MTR |
| [24] | - | • | 44 | - | - | - | 44 | MTR |

| | | | | | | | | |
|---|---|---|---|---|---|---|---|---|
| 25 | - | • | 12 | 12 | - | - | 12 | MTR |
| 26 | - | • | 36 | - | 24 | - | 23 | MTR |
| 27 | | • | 10 | 5 | | - | 5 | MTR |
| 28 | • | - | - | - | - | 18 (CIS) | 18 | MTR |
| 29 | • | - | 11 | - | - | 36 (CIS) | - | MTR |
| 30 | • | - | - | - | - | 100 (CIS) | 50 | MTR |
| 31 | - | • | 20 | - | - | 7 (chronic progressive) | 10 | MTR |
| 32 | - | • | 7 | 7 | - | - | 5 | MTR |
| 33 | - | • | 39 | 19 | 9 | 9 (benign); 20 (CIS) | 20 | MTR |
| 34 | • | - | 34 (32) | 19 (16) | - | 20 (19) [CIS] | - | MTR |
| 35 | - | • | 31 | 10 | - | 28 (CIS) | 19 | MTR |
| 36 | • | - | - | - | - | 43 (CIS) | 22 | MTR |
| 37 | - | • | 16 | 11 | - | - | 16 | MTR |
| 38 | - | • | 10 | - | - | 25 (benign) | 10 | MTR |
| 39 | • | - | - | - | - | 12 (CIS & RRMS) | 12 | MTR |
| 40 | - | • | 66 | 30 | 21 | - | 26 | MTR |
| 41 | - | • | 40 | 24 | 14 | - | - | MTR |
| 42 | - | • | 66 | 30 | 21 | - | 26 | MTR |
| 43 | - | • | 10 | 1 | | - | - | MTR |
| 44 | • | - | - | - | - | 24 (CIS) | 20 | MTR |
| 45 | - | • | 69 | 32 | - | - | 41 | MTR |
| 46 | - | • | 17 | 12 | 3 | - | 17 | MTR |
| 47 | - | • | 16 | 26 | 26 | 11 (benign) | 23 | MTR |
| 48 | - | • | 22 | 32 | 32 | - | - | MTR |
| 49 | - | • | 80 | - | - | 44 (CIS) | - | MTR |
| 50 | • | - | 38 | 13 | 1 | 1 (benign) | - | MTR; (MWF) |
| 51 | • | - | 7 | 1 | 1 | - | | MTR |
| 52 | • | - | 83 | - | 22 | 27 | - | MTR; (MWF) |
| 53 | • | 105 [29] | - | 26 [0] | - | 19 | - | MTR |
| 54 | - | • | 43 | 28 | - | - | 38 | MTR |
| 55 | | • | 14 | 9 | | - | 9 | MTR |
| 56 | • | - | 13 | 4 | | - | - | MTR |

| | | | | | | | | |
|---|---|---|---|---|---|---|---|---|
| 57 | • | - | 14 | 7 | 14 | - | 36 | MTsat |
| 58 | - | • | 15 | 21 | | - | 36 | MTsat |
| 59 | - | • | 51 | 28 | 19 | - | 29 | MTR |
| 60 | - | • | 26 | 12 | | - | - | MTR |
| 61 | • | - | 16 | 2 | 1 | - | - | MTR |
| 62 | • | - | 27 | 14 | 6 | 21 | - | MTR |
| 63 | - | • | 22 | 8 | 6 | - | 18 | MTR |
| 64 | - | • | 66 | 36 | | - | 11 | MTR |
| 65 | • | - | 35 | 20 | 14 | 29 | - | MTR |
| 66 | • | - | 8 | 3 | 2 | - | - | MTR |
| 67 | - | • | 44 | 27 | - | - | 22 | MTR |
| 68 | • | - | 32 | 2 | - | 19 (CIS); 8 | - | MTR |
| 69 | • | - | 20 | 10 | | - | 8 | MTR |
| 70 | - | • | 11 | 14 | 5 | - | - | MTR |
| 71 | - | • | 11 | 14 | 5 | - | 12 | MTR |
| 72 | • | - | 47 | 5 | - | 17 (CIS) | - | MTR |
| 73 | • | - | 3 | - | - | 3 (benign) | 5 | qMT including $f$,$T2_b$ |
| 74 | - | • | 2 | - | 1 | - | 3 | MTR |
| 75 | • | - | 40 | 24 | 14 | - | - | MTR |
| 76 | • | - | 50 | 24 | 14 | - | 27 | MTR |
| 77 | - | • | 52 | 24 | 14 | - | 29 | MTR |
| 78 | • | - | 43 | 22 | 10 | - | 29 | MTR |
| 79 | - | • | 26 | 13 | 8 | - | 29 | MTR |
| 80 | - | • | 7 | 7 | - | - | 5 | MTR |
| 81 | - | • | 34 | 19 | - | 20 (CIS) | 13 | MTR |
| 82 | - | • | 161 | | - | - | - | MTR |
| 83 | - | • | 34 | 19 | - | 20 (CIS) | 16 | MTR |
| 84 | • | - | 25 | 17 | - | - | - | MTR |
| 85 | - | • | 40 | 28 | 9 | - | - | MTR |
| 86 | • | - | 10 | 15 | 5 | - | - | MTR |
| 87 | - | • | 39 | - | 25 | - | 20 | MTR |
| 88 | - | • | 44 | 25 | 19 | - | 35 | MTR |

| | | | | | | | | |
|---|---|---|---|---|---|---|---|---|
| 89 | - | ● | 31 | 14 | 16 | - | 32 | MTR |
| 90 | - | ● | 50 | 10 | - | - | 20 | MTR |
| 91 | ● | - | 1 | 2 | - | - | - | MTR |
| 92 | ● | - | 477 | 222 | 30 | 29 (CIS) | - | MTR |
| 93 | - | ● | 28 | 17 | - | - | 19 | MTR |
| 94 | - | ● | 33 | 20 | 13 | 11 (benign) | 20 | MTR |
| 95 | ● | - | 14 | 2 | 2 | 2 (benign) | 7 | MTR; qMT inc. $f$, $T2_b$ |
| 96 | ● | - | 10 | 5 | 4 | - | 9 | qMT inc. $T2_b$, $f$ |
| 97 | ● | - | 32 | - | 3 | 38 (CIS) | 23 | MTR |
| 98 | - | ● | 70 | 25 | - | - | 63 | MTR |
| 99 | ● | - | 11 | 4 | | - | 11 | MTR |
| 100 | ● | - | 28 | 16 | | - | - | MTR |
| 101 | ● | - | 6 | 5 | | - | 11 | MTR |
| 102 | ● | - | 8 | 11 | 1 | - | 5 | SNR & CNR with/without MT |
| 99 | - | ● | 11 | 4 | | - | - | MTR |
| 103 | ● | - | 5 | 1 | 1 | - | 7 | MTR |
| 104 | ● | - | 35 | 19 | 12 | - | 23 | MTR |
| 105 | - | ● | 5 | 4 | - | - | 10 | MTR; MWF |
| 106 | - | ● | 34 | 18 | 11 | - | 22 | MTR |
| 107 | - | ● | 35 | 19 | 12 | - | 23 | MTR |
| 108 | ● | - | 3 | 2 | 1 | - | 5 | MTR |
| 109 | ● | - | 145 | | | - | - | MTR |
| 110 | ● | - | 42 | 3 | | - | - | MTR |
| 111 | - | ● | 46 | 26 | - | - | 36 | MTR |
| 112 | ● | - | 30 | 30 | 25 | - | 36 | MTR |
| 113 | ● | - | 30 | 30 | 25 | - | - | MTR |
| 114 | - | ● | 18 | 12 | - | - | 14 | MTR; R1; MPF |
| 115 | - | ● | 32 | 17 | - | 10 (CIS) | 14 | MTR |
| 116 | ● | - | 27 | 12 | 4 | - | 20 | MTR |
| 117 | ● | - | 554 | 227 | 34 | - | - | MTR |

*Supplementary Table 2: Overview of studies excluded from final analysis as relapse-onset multiple sclerosis (MS), not solely relapsing-remitting MS. MT: magnetisation transfer; MTR: MT ratio; MTsat: MTsat; MWF: myelin water fraction; PSR: pool size ratio; qMT: quantitative MT; RRMS: relapsing-remitting MS; SPMS: secondary progressive MS.*

| Citation | SPMS only | RRMS (n) | SPMS (n) | Healthy (n) | MT Technique |
|---|---|---|---|---|---|
| [1] | - | 27 | 3 | 30 | MTR |
| [2] | ● | - | 1 | 6 | qMT inc. $T2_b$, PSR |
| [3] | - | 20 | 4 | 24 | MTR |
| [4] | - | 21 | 8 | 10 | MTR |
| [5] | - | 8 | 2 | 12 [8] | MTR |
| [6] | - | 27 | 5 | - | MTR |
| [7] | - | 19 | 11 | 15 | MTR |
| [8] | - | 17 | 14 | 14 | MTR |
| [9] | ● | - | 1 | - | MTR |
| [10] | - | 15 | 5 | - | MTR; CNR |
| [11] | ● | - | 72 | - | MTR |
| [12] | - | 8 | 11 | 20 | MTR |
| [13] | - | 13 | 6 | - | MTR |
| [14] | - | 36 | 19 | - | MTR |
| [15] | ● | - | 117 | - | MTR |
| [16] | ● | - | 1 | - | MTR |
| [17] | - | 9 | 6 | - | MTR |
| [18] | ● | - | 118 | - | MTR |
| [19] | ● | - | 117 | - | MTR |
| [20] | - | 46 | | - | MTR |
| [21] | - | 44 | 28 | 20 | MTR |

| | | | | | |
|---|---|---|---|---|---|
| [22] | • | - | 82 | - | MTR |
| [23] | - | 45 | 7 | 20 | MTR |
| [24] | - | 14 (1 benign) | 5 | - | MTR; (MWF) |
| [25] | - | 128 | 6 | - | MTR; MTsat |
| [26] | - | 5 | 5 | - | F; MTR; $PD_f$ |
| [27] | - | 21 | 2 | - | MTR |
| [28] | - | 10 | 10 | - | MTR |
| [29] | - | 10/13/NA | 10/7/67 | - | MTR |
| [30] | - | 4 | 4 | 5 | F |
| [31] | - | 36 | 2 | 11 | MTR |
| [32] | - | 11 | 3 | - | MTR |
| [33] | - | 41 | 26 | 30 | MTR |
| [34] | - | 9 | 7 | - | MTR |
| [35] | | 161 | | - | MTR |
| [36] | - | 8 | 14 | - | MTR |
| [37] | - | 21 | 4 | 12 | MTR |
| [38] | - | 8 | 10 | 12 | MTR |
| [39] | - | 12 | 8 | 10 | MTR |
| [40] | - | 1 | 2 | - | MTR |
| [41] | - | 30 (29) | - (1) | - | MTR |
| [42] | - | 5 | 5 | - | MTR |
| [43] | - | 8 | 33 | - | MTR |
| [44] | - | 16 | 5 | - | MT with contrast ratio |
| [45] | - | 8 | 3 | - | MTR |
| [46] | - | 211 | 74 | - | MTR |
| [47] | - | 73 | 30 | 22 | MTR |

**Reference for** Error! Reference source not found.

*Supplementary Table 3: Study sample characteristics (n=86) for relapsing-remitting multiple sclerosis patients and controls. See main text for reference list. *Numbers for recruitment and analysis were identical with the exception of those studies marked. RRMS: relapsing-remitting multiple sclerosis; F:M: female-to-male; DMDs: disease-modifying drugs; wks: weeks; mths: months; yrs: years; IfN: interferon; PegIfN: pegylated interferon; i.v.: intraveneous; s.c.: subcutaneous. ♣ median age, mean not reported*

| | | RRMS | | | | | Healthy Controls | | | |
|---|---|---|---|---|---|---|---|---|---|---|
| | | Recruited | | Analysed | | | Analysed* | | | |
| **Citation** | **Location of Study Centre(s)** | **n** | **F:M** | **n** | **F:M** | **Mean Age (in yrs)** | **n** | **F:M** | **Mean Age (in yrs)** | **DMDs & Steroids** |
| Al-Radaideh, Athamneh, Alabadi, Hbahbih [24] | Zarqa, Jordan | 30 | 1.31 | 30 | 1.31 | 31.27 | 30 | 1.14 | 32.35 | No corticosteroid treatment for 4 wks preceeding study |
| Amann, Sprenger, Naegelin, Reinhardt, Kuster, Hirsch, Kappos, Radue, Stippich, Bieri [25] | Basel | 31 | 1.58 | 27 | 1.58 | 54.4 | - | - | - | All unknown DMD; no corticosteroid 3mths prior |
| Arnold, Gold, Kappos, Bar-Or, Giovannoni, Selmaj, Yang, Zhang, Stephan, Sheikh, Dawson [26] | International | 540 | 3.39 | 392 | - | 38.4 | - | - | - | Delayed-release dimethyl fumarate (i.e. BG-12, 240mg, 3 OR 2 times daily) OR placebo |
| Arnold, Calabresi, Kieseier, Liu, You, Fiore, Hung [27] | International | 1512 | 2.43 | 858 | - | 36.5 | - | - | - | PegIfN-β1a (s.c., 125µg every 2wks) OR PegIfN-β1a (s.c., 125µg every 4wks) OR placebo (then |

| | | | | | | | | | | |
|---|---|---|---|---|---|---|---|---|---|---|
| | | | | | | | | | | treat as above at 48wks) |
| Audoin, Davies, Rashid, Fisniku, Thompson, Miller [28] | London/ Marseille | 38 | 2.80 | 38 | 2.80 | 36.3 | 45 | 1.37 | 34 | Unknown |
| Bellmann-Strobl, Stiepani, Wuerfel, Bohner, Paul, Warmuth, Aktas, Wandinger, Zipp, Klingebiel [29] | Berlin | 17 | 1.43 | 6 | - | 33 | 17 | 1.43 | 30.5 | IfN-β1a (s.c., 22μg, 3 times a week) |
| Bernitsas, Kopinsky, Lichtman-Mikol, Razmjou, Santiago-Martinez, Yarraguntla, Bao [30] | Detroit, MI | 50 | - | 44 | 1.75 | 40.8 | 27 | 2.38 | 37.2 | daily .5mg fingolimod with 30 day washout period for IfN-β /glatirmaer acetate; steroid-free for at least 1 month. No monoclonal antibodies (e.g. alemtuzumab, natalizumab, rituximab, daclizumab) |
| Bonnier, Roche, Romascano, Simioni, Meskaldji, Rotzinger, Lin, Menegaz, Schluep, Du Pasquier, Sumpf, | Lausanne | 36 | 2.00 | 36 | 2.00 | 34.8 | 18 | 1.00 | 33 | 30 IfN-β or fingolimod for 3+ months |

| | | | | | | | | | | |
|---|---|---|---|---|---|---|---|---|---|---|
| Frahm, Thiran, Krueger, Granziera [31] | | | | | | | | | | |
| Bonnier, Roche, Romascano, Simioni, Meskaldji, Rotzinger, Lin, Menegaz, Schluep, Du Pasquier, Sumpf, Frahm, Thiran, Krueger, Granziera [32] | Lausanne | 36 | 2.00 | 36 | 2.00 | 34.8 | 18 | 1.00 | 33 | 30 IfN-β or fingolimod for 3+ months |
| Bonnier, Marechal, Fartaria, Falkowskiy, Marques, Simioni, Schluep, Du Pasquier, Thiran, Krueger, Granziera [33] | Lausanne | 23 | 1.88 | 23 | 1.88 | 35.7 | 9 | 1.25 | 34.3 | 20 IfN-β or fingolimod for 3+ mths (22 at 2yrs); no corticosteroids 3mths prior |
| Bonnier, Fischi-Gomez, Roche, Hilbert, Kober, Krueger, Granziera [34] | Lausanne | 15 | 2.00 | 13 | - | 32 | 4 | 1.50 | 39.56 | No corticosteroid therapy for at least 3 mth prior. High dosage of either IfN-β or fingolimod |
| Catalaa, Grossman, Kolson, Udupa, Nyul, Wei, Zhang, Polansky, Mannon, McGowan [35] | Pennsylvania | 23 | 4.75 | 23 | 6.33 | - | 25 | - | - | No immunosuppressant/ immunomodulatory drugs at time of MRI, except for corticosteroids for exacerbations |

| | | | | | | | | | |
|---|---|---|---|---|---|---|---|---|---|
| Cercignani, Iannucci, Rocca, Comi, Horsfield, Filippi [36] | Milan | 35 | 1.33 | 35 | 1.33 | 28 | 24 | 1.67 | 29 | No immunomodulatory/ immunosuppress antdrugs for 1yr prior and during study; no steroid for 3mths prior |
| Cercignani, Basile, Spano, Comanducci, Fasano, Caltagirone, Nocentini, Bozzali [37] | Rome | 13 | 1.60 | 13 | 1.60 | 39.4 | 14 | 0.75 | 34.9 | 9 IfN-β; 4 glatiramer acetate; no steroids 3 mth prior |
| Codella, Rocca, Colombo, Martinelli-Boneschi, Comi, Filippi [38] | Milan | 28 | 2.11 | 28 | 2.11 | 37.6/39.1 | 30 | 1.25 | 38.6 | Untreated |
| Colasanti, Guo, Muhlert, Giannetti, Onega, Newbould, Ciccarelli, Rison, Thomas, Nicholas, Muraro, Malik, Owen, Piccini, Gunn, Rabiner, Matthews [39] | London | 11 | 10.00 | 11 | 10.00 | 45.1 | 11 | 0.83 | 45.7 | 2 natalizumab; 6 IfN-β; 3 none |
| Cronin, Xu, Bagnato, Gochberg, Gore, Dortch [40] | Nashville | 2 | - | 2 | - | - | 8 | 0.14 | 29.9 | Unknown |
| Davies, Ramani, Dalton, Tozer, Wheeler-Kingshott, Barker, | London | 5 | 1.50 | 5 | 1.50 | 40 | 5 | 1.50 | 38 | Unknown |

| | | | | | | | | | | |
|---|---|---|---|---|---|---|---|---|---|---|
| Thompson, Miller, Tofts [41] | | | | | | | | | | |
| Davies, Ramio-Torrenta, Hadjiprocopis, Chard, Griffin, Rashid, Barker, Kapoor, Thompson, Miller [42] | London | 38 | 2.80 | 38 | 2.80 | 36.3 | 35 | 1.19 | 38.5 | No DMT at time of imaging |
| Davies, Altmann, Hadjiprocopis, Rashid, Chard, Griffin, Tofts, Barker, Kapoor, Thompson, Miller [43] | London | 23 | 4.75 | 21 | - | 37 | 19* | 1.11 | 34 | Untreated at baseline; 7 on IfN-β by 1yr |
| Davies, Altmann, Rashid, Chard, Griffin, Barker, Kapoor, Thompson, Miller [44] | London | 23 | 4.75 | 22 | - | 37 | 19* | 1.11 | 34 | Untreated at baseline; 7 on IfN-β by 1yr |
| Deloire, Ruet, Hamel, Bonnet, Dousset, Brochet [45] | Bordeaux | 44 | 3.40 | 44 | 3.40 | 39 | 56 | 1.80 | 38.2 | 95.6% DMDs |
| De Stefano, Narayanan, Francis, Smith, Mortilla, Tartaglia, Bartolozzi, Guidi, Federico, Arnold [46] | Montreal/ Siena | 60 | 2.16 | 57 | 2.16 | 35 | 42 | 1.48 | 35 | None for 1 mth prior |
| Dortch, Li, Gochberg, Welch, Dula, Tamhane, Gore, Smith [47] | Nashville | 2 | 1.00 | 2 | 1.00 | - | 9 | 2.00 | - | Unknown |
| Dortch, Bagnato, Gochberg, Gore, Smith [48] | Nashville | 1 | - | 1 | - | 37 | 4 | 1.00 | - | Unknown |

| | | | | | | | | | | |
|---|---|---|---|---|---|---|---|---|---|---|
| Ernst, Chang, Walot, Huff [49] | Torrance, CA | 1 | - | 1 | - | 20 | - | - | - | IfN-β1b, 0.25mg every other day; 3 day course of methylprednisolone (1 g/d, i.v.) |
| Fatemidokht, Harirchian, Faghihzadeh, Tafakhori, Oghabian [50] | Tehran | 18 | 0.50 | 18 | 0.50 | 37 ♣ | - | - | - | Unknown |
| Fazekas, Ropele, Enzinger, Seifert, Strasser-Fuchs [51] | Graz | 12 | 2.00 | 12 | 2.00 | - | - | - | - | Oral IfN-β1a OR placebo |
| Filippi, Rocca, Rizzo, Horsfield, Rovaris, Minicucci, Colombo, Comi [52] | Milan | 10 | 2.33 | 10 | 2.33 | 30.4 | - | - | - | Methylprednisolone for relapses but >10 days prior to MRI; no other immunomodulatory or immunosuppressants |
| Filippi, Rocca, Sormani, Pereira, Comi [53] | Milan | 10 | 2.33 | 10 | 2.33 | 30.4 | - | - | - | Methylprednisolone for relapses but >10 days prior to MRI; no other immunomodulatory or immunosuppressants |

| | | | | | | | | | | |
|---|---|---|---|---|---|---|---|---|---|---|
| Filippi, Rocca, Pagani, De Stefano, Jeffery, Kappos, Montalban, Boyko, Comi, Group [54] | International | 92 | - | 63 | - | - | - | - | - | 38 (31 at 24mths) laquinomod; 37 (32 at 24 mths) placebo |
| Fooladi, Sharini, Masjoodi, Khodamoradi [55] | Tehran | 30 | 1.73 | 30 | 1.73 | 30.2 | 30 | 2.33 | 30 | Unknown |
| Fooladi, Riyahi Alam, Sharini, Firouznia, Shakiba, Harirchian [56] | Tehran | 12 | 2.00 | 12 | 2.00 | 31 | 12 | 3.00 | 29.5 | Immunomodulatory therapy |
| Fritz, Keller, Calabresi, Zackowski [57] | Baltimore | 30 | - | 29 | 1.42 | 48.69 | 29 | 2.50 | 50.76 | No corticosteroids for 30 days prior to testing |
| Frohman, Dwyer, Frohman, Cox, Salter, Greenberg, Hussein, Conger, Calabresi, Balcer, Zivadinov [58] | Buffalo/ Dallas | 12 | 1.40 | 12 | 1.40 | 40.6 | 4 | 1.00 | 35.3 | None for 1 month prior |
| Ge, Grossman, Udupa, Babb, Kolson, McGowan [59] | Philadelphia | 18 | 5.00 | 18 | 5.00 | 34.1 | 18 | 2.00 | 33.9 | Unknown |
| Ge, Grossman, Babb, He, Mannon [60] | New York | 22 | 2.67 | 22 | 2.67 | 35.2 | - | - | - | No immunomodulatory treatment previously but short course of steroids for relapses |

| | | | | | | | | | |
|---|---|---|---|---|---|---|---|---|---|
| Giacomini, Levesque, Ribeiro, Narayanan, Francis, Pike, Arnold [61] | Montreal | 6 | - | 6 | - | 43.7 | - | - | - | 1 untreated; 1 natalizumab; 2 glatiramer acetate; 1 IfN-βa (s.c.); 1 IfN-βb |
| Goodkin, Rooney, Sloan, Bacchetti, Gee, Vermathen, Waubant, Abundo, Majumdar, Nelson, Weiner [62] | San Francisco | 22 | 1.00 | 11 | - | 34.3 | 11 | 0.83 | 38.1 | No immunomodulatory therapy or immunosuppressantdrugs; MRI >14 days after steroids |
| Gracien, Jurcoane, Wagner, Reitz, Mayer, Volz, Hof, Fleischer, Droby, Steinmetz, Groppa, Hattingen, Deichmann, Klein [63] | Frankfurt | 22 | 4.50 | 22 | 4.50 | 34.7 | 10 | 4.00 | 34.2 | 3 natalizumab; 1 fingolimod; 1 dimethyl fumarate; 1 glatiramer acetate; 14 untreated; 2 unknown |
| Griffin, Chard, Parker, Barker, Thompson, Miller [64] | London | 22 | 2.14 | 22 | 2.14 | 36.6 | 11 | 1.75 | 37 | No DMDs; no steroids for 1 mth prior |
| Guo, Jewells, Provenzale [65] | Durham | 12 | 2.00 | 12 | 2.00 | 39 | - | - | - | Unknown |
| Helms, Dathe, Kallenberg, Dechent [7] | Gottingen | 1 | - | 1 | - | 27 | 7 | - | - | Unknown |
| Iannucci, Rovaris, Giacomotti, Comi, Filippi [66] | Milan | 34 | 1.62 | 34 | 1.62 | 34.8 | 15 | 1.50 | 34 | Unknown |

| | | | | | | | | | | |
|---|---|---|---|---|---|---|---|---|---|---|
| Kamagata, Zalesky, Yokoyama, Andica, Hagiwara, Shimoji, Kumamaru, Takemura, Hoshino, Kamiya, Hori, Pantelis, Hattori, Aoki [67] | Tokyo | 14 | - | 14 | - | 42.8 | 14 | 14.0 | 43.2 | Unknown |
| Karampekios, Papanikolaou, Papadaki, Maris, Uffman, Spilioti, Plaitakis, Gourtsoyiannis [68] | Crete | 12 | 1.40 | 12 | 1.40 | 27.2 | 5 | 1.50 | 27.6 | Unknown |
| Kita, Goodkin, Bacchetti, Waubant, Nelson, Majumdar [69] | San Francisco | 22 | 1.00 | 8 | - | 38 | - | - | - | IfN-β1a; no methylprednisolone 14 days prior to MRI |
| Kuhle, Barro, Disanto, Mathias, Soneson, Bonnier, Yaldizli, Regeniter, Derfuss, Canales, Schluep, Du Pasquier, Krueger, Granziera [70] | Lausanne | 31 | 1.82 | 19 | - | 32 | 18 | 1.25 | 31 | Untreated at baseline; at follow-up, 11 IfN-β1a; 1 IfN-β1b; 4 glatiramer acetate; 6 fingolimod |
| Levesque, Giacomini, Narayanan, Ribeiro, Sled, Arnold, Pike [71] | Montreal | 5 | - | 5 | - | 42.6 | 5 | 0.67 | - | 1 glatiramer acetate; 4 untreated |
| Lin, Tench, Morgan, Constantinescu [72] | Nottingham | 36 | 2.60 | 36 | 2.60 | 37.5 | 13 | 1.60 | 34 | Unknown |
| Mangia, Carpenter, Tyan, Eberly, Garwood, Michaeli [73] | Minneapolis | 9 | 3.50 | 9 | 3.50 | 38 | 7 | 2.50 | 37 | Unknown |

| | | | | | | | | | | |
|---|---|---|---|---|---|---|---|---|---|---|
| McKeithan, Lyttle, Box, O'Grady, Dortch, Conrad, Thompson, Rogers, Newhouse, Pawate, Bagnato, Smith [74] | Nashville | 19 | 3.75 | 19 | 3.75 | 38 | 37 | 2.08 | 32 | Unknown |
| Mesaros, Rocca, Sormani, Valsasina, Markowitz, De Stefano, Montalban, Barkhof, Ranjeva, Sailer, Kappos, Comi, Filippi [75] | International | 42 | 1.80 | 42 | 1.80 | 36.7 | - | - | - | Placebo |
| Miller, Fox, Phillips, Hutchinson, Havrdova, Kita, Wheeler-Kingshott, Tozer, MacManus, Yousry, Goodsell, Yang, Zhang, Viglietta, Dawson, investigators [76] | International | 758 | 1.71 | 555 | - | 37.5 | - | - | - | Delayed-release dimethyl fumarate (240mg 2 or 3 times daily) OR glatiramer acetate (20mg, daily) OR placebo |
| Muhlert, Atzori, De Vita, Thomas, Samson, Wheeler-Kingshott, Geurts, Miller, Thompson, Ciccarelli [77] | London | 18 | 0.50 | 14 | - | 43.5 | 17 | 0.55 | 39.7 | No corticosteroids fro 4 wks prior; treatment unknown |
| O'Muircheartaigh, Vavasour, Ljungberg, Li, Rauscher, Levesque, Garren, Clayton, Tam, Traboulsee, Kolind [78] | Vancouver | 56 | 1.95 | 24 | 1.67 | 37 | 38 | 1.92 | 35 | Ocrelizumab or IfN-β1a |

| | | | | | | | | | | |
|---|---|---|---|---|---|---|---|---|---|---|
| Oreja-Guevara, Charil, Caputo, Cavarretta, Sormani, Filippi [79] | Milan | 22 | 2.14 | 22 | 2.14 | 36.6 | - | - | - | Untreated |
| Ostuni, Richert, Lewis, Frank [80] | Bethesda | 9 | 3.50 | 9 | 3.50 | 37 | 5 | 0.67 | 37 | Unknown |
| Patel, Grossman, Phillips, Udupa, McGowan, Miki, Wei, Polansky, van Buchem, Kolson [81] | Philadelphia | 20 | 4.00 | 20 | 4.00 | 37 | - | - | - | 8 steroid treatment ~1mth for exacerbation; 3 received IfN-α |
| Preziosa, Pagani, Moiola, Rodegher, Filippi, Rocca [82] | Milan | 104 | - | 52 | 1.36 | 36.85 | - | - | - | 24 fingolimod; 28 natalizumab |
| Reich, White, Cortese, Vuolo, Shea, Collins, Petkau [83] | Bethesda | 12 | 0.33 | 6 | 0.20 | 35 | - | - | - | IfN-β; 2 glatiramer acetate; 8 untreated |
| Reitz, Hof, Fleischer, Brodski, Groger, Gracien, Droby, Steinmetz, Ziemann, Zipp, Deichmann, Klein [84] | Frankfurt | 9 | 2.00 | 9 | 2.00 | - | 12 | 0.71 | - | 3 IfN-β1a; 1 glatiramer acetate; 2 natalizumab; 1 fingolimod; 2 dimethyl fumarate |
| Richert, Ostuni, Bash, Duyn, McFarland, Frank [85] | Bethesda | 8 | 1.00 | 8 | 1.00 | 37 | 5 | 1.00 | 37.2 | IfN-β1b 8mU, s.c., every other day |
| Richert, Ostuni, Bash, Leist, McFarland, Frank [86] | Bethesda | 4 | 1.00 | 4 | 1.00 | 29 | - | - | - | IfN-β1a; methylprednidolo |

| | | | | | | | | | | |
|---|---|---|---|---|---|---|---|---|---|---|
| | | | | | | | | | | ne (corticosteroid i.v.) |
| Rocca, Falini, Colombo, Scotti, Comi, Filippi [87] | Milan | 14 | 1.33 | 14 | 1.33 | 37.6 | 15 | 1.50 | 38.6 | No steroid for 6 mths prior; never treated with immunomodulatory drugs or immunosuppressants |
| Romascano, Meskaldji, Bonnier, Simioni, Rotzinger, Lin, Menegaz, Roche, Schluep, Pasquier, Richiardi, Van De Ville, Daducci, Sumpf, Fraham, Thiran, Krueger, Granziera [88] | Lausanne | 28 | 1.80 | 28 | 1.80 | 34.32 | 16 | 1.29 | 33.06 | 28 IfN-β or fingolimod for 3+ months |
| Ropele, Strasser-Fuchs, Augustin, Stollberger, Enzinger, Hartung, Fazekas [89] | Graz | 9 | 2.00 | 9 | 2.00 | 38 | 8 | 1.00 | - | 5 on long-term immunomodulatory therapy |
| Rovira, Alonso, Cucurella, Nos, Tintore, Pedraza, Rio, Montalban [90] | Barcelona | 11 | 0.22 | 11 | 0.22 | 32 | 6 | - | - | No prior cytotoxic or immunomodulatory therapy; no corticosteroids during study |
| Rudick, Lee, Simon, Fisher [91] | Cleveland | 30 | - | 30 | - | 36.3 | - | - | - | Initially placebo or IfN-β1a; DMTs changed over |

| | | | | | | | | | | |
|---|---|---|---|---|---|---|---|---|---|---|
| | | | | | | | | | | time but also untreated periods |
| Saccenti, Hagiwara, Andica, Yokoyama, Fujita, Kato, Maekawa, Kamagata, Le Berre, Hori, Wada, Tateishi, Hattori, Aoki [92] | Tokyo | 37 | - | 21 | 9.50 | 37.9 | - | - | - | Unknown |
| Schwartz, Tagge, Powers, Ahn, Bakshi, Calabresi, Constable, Grinstead, Henry, Nair, Papinutto, Pelletier, Shinohara, Oh, Reich, Sicotte, Rooney, Chu, Tauhid, Tummala, Kober, Pan, Sati, Simon, Stern, Cooperative [93] | Oregon (7 sites) | 1 | 0.00 | 1 | 0.00 | 45 | - | - | - | Dimethyl fumarate |
| Siemonsen, Young, Bester, Sedlacik, Heesen, Fiehler, Stellmann [94] | Hamburg | 23 | 1.30 | 23 | 1.30 | 41 | - | - | - | Unknown |
| Sled, Pike [95] | Montreal/Siena | 1 | - | 1 | - | - | 2 | - | - | Unknown |
| Smith, Farrell, Jones, Reich, Calabresi, van Zijl [96] | Baltimore | 5 | - | 5 | - | - | 9 | - | 30 | Unknown |
| Thaler, Faizy, Sedlacik, Bester, Stellmann, Heesen, Fiehler, Siemonsen [97] | Hamburg | 35 | 1.69 | 35 | 1.69 | 37 | - | - | - | Dimethyl fumarate |

| | | | | | | | | | | |
|---|---|---|---|---|---|---|---|---|---|---|
| van den Elskamp, Knol, Vrenken, Karas, Meijerman, Filippi, Kappos, Fazekas, Wagner, Pohl, Sandbrink, Polman, Uitdehaag, Barkhof [98] | International | 32 | 1.67 | 24 | - | 32.3 | - | - | - | Oral IfN-β1a OR placebo |
| Van Obberghen, McHinda, le Troter, Prevost, Viout, Guye, Varma, Alsop, Ranjeva, Pelletier, Girard, Duhamel [99] | Marseille | 25 | 4.00 | 25 | 4.00 | 41.5 | 20 | 2.33 | 40.5 | 10 DMTs |
| Weinstock-Guttman, Zivadinov, Tamano-Blanco, Abdelrahman, Badgett, Durfee, Hussein, Feichter, Patrick, Benedict, Ramanathan [100] | Buffalo | 52 | 3.73 | 39 | 4.57 | 49.6 | - | - | - | Unknown (maybe IfN-β1a), no steroid for 4wks prior |
| Yarnykh [101] | Seattle | 2 | - | 2 | - | 42.5 | 2 | 2.00 | 47 | Unknown |
| Yiannakas, Tozer, Schmierer, Chard, Anderson, Altmann, Miller, Wheeler-Kingshott [102] | London | 10 | 2.33 | 10 | 2.33 | 53 | - | - | - | Unknown |
| Zhang, Wen, Chen, Tian, Xue, Ren, Li, Fan, Ren [103] | Baoji, Shaanxi | 18 | 2.00 | 18 | 2.00 | 31.2 | 16 | 1.67 | 30.4 | Unknown |

| | | | | | | | | | | |
|---|---|---|---|---|---|---|---|---|---|---|
| Zhou, Zhu, Grimaud, Hermier, Rovaris, Filippi [104] | Lyon/Chatres/ Milan | 10 | 1.50 | 10 | 1.50 | 38.5 | 10 | 1.33 | 31.4 | No steroid treatment for 3 mths prior |
| Zivadinov, De Masi, Nasuelli, Bragadin, Ukmar, Pozzi-Mucelli, Grop, Cazzato, Zorzon [105] | Trieste | 63 | 2.15 | 63 | 2.15 | 35.4 | 30 | - | - | No immunomodulatory drugs/steroids 3 mths prior |
| Zivadinov, Hussein, Stosic, Durfee, Cox, Cookfair, Hashmi, Abdelrahman, Garg, Dwyer, Weinstock-Guttman [106] | Buffalo | 19 | 2.80 | 13 | - | 42.3 | 16 | 15.0 | 44.2 | Glatiramer acetate (20mg/day sc) |
| Zivadinov, Ramanathan, Ambrus, Hussein, Ramasamy, Dwyer, Bergsland, Minagar, Weinstock-Guttman [107] | Buffalo | 47 | 3.27 | 47 | 3.27 | 45.5 | - | - | - | IfN-β1a (i.m., 30μg) |
| Zivadinov, Dwyer, Markovic-Plese, Kennedy, Bergsland, Ramasamy, Durfee, Hojnacki, Hayward, Dangond, Weinstock-Guttman [108] | Buffalo | 21 | 2.00 | 21 | 2.00 | 39.9 | 15 | 1.14 | 36.7 | IfN-β1a (s.c., 44μg) |

*Supplementary Table 4: Overview of usage of disease-modifying drugs (DMDs) across studies. Reference numbers refer to main text references.*

| Number of DMDs | | n | % | Citation |
|---|---|---|---|---|
| 0 | No DMDs | 16 | 18.60 | 30,37,38,40,44,48,53,55,57,71,74,80,86-88,91 |
| | Placebo only | 1 | 1.16 | 90 |
| 1 | 1 (or untreated) | 4 | 4.65 | 45,46,62,92 |
| | 1 (all treated) | 11 | 12.79 | 25,26,31,32,70,81,82,95,98,101,103 |
| >1 | No placebo | 14 | 16.28 | 33-36,39,41,56,61,67,69,72,93,94,106 |
| | With placebo | 7 | 8.14 | 28,85,102,104,105,107,108 |
| Unknown | Unspecified | 5 | 5.81 | 47,51,73,76,83 |
| Data missing | | 28 | 32.56 | 7,24,27,29,42,43,49,50,52,54,58-60,63-66,68,75,77-79,84,89,96,97,99,100 |

*DMDs: disease-modifying drugs.*

*Supplementary Figure 1: Predicted longitudinal evolution of magnetisation transfer ratio (MTR) over three years across different brain regions of patients with relapsing-remitting multiple sclerosis, modelled from longitudinal papers with reported means and variance. Marginal mean estimates were calculated from a linear regression model with mean MTR as the dependent variable, timepoint and brain region as fixed effects, and study as a random effect. Further sub-groupings per study were added as an additional nested factor within study (e.g. placebo versus treatment groups, active versus reactive lesions).*

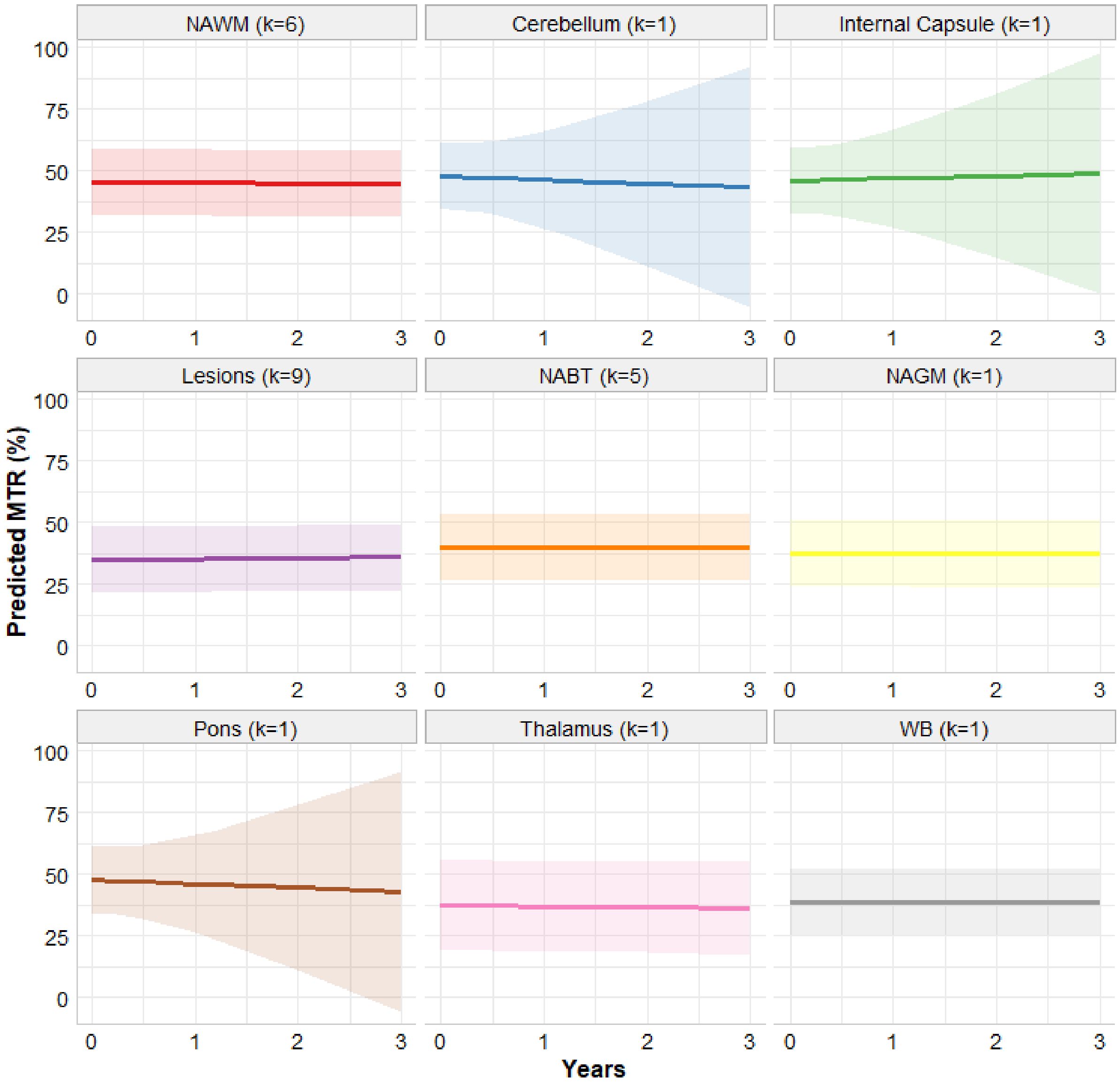

*Supplementary Table 5: Linear mixed model fit results examining longitudinal change in mean MTR across brain sub-regions*

| **Linear Mixed Model Fit by Maximum Likelihood (*lmer* package in RStudio)** | | | | | | |
|---|---|---|---|---|---|---|
| **Random effects:** | | | | | | |
| | | | **Variance** | | **SD** | |
| Study/Subgroup | (intercept) | | 4.64 | | 2.15 | |
| Study | (intercept) | | 38.41 | | 6.20 | |
| Residual | | | 2.41 | | 1.55 | |
| **Fixed effects:** | | | | | | |
| ***Predictors*** | ***Estimates*** | ***CI*** | | ***df*** | ***t-value*** | ***p*** |
| (intercept) | 45.11 | 41.10 | 49.13 | 13.88 | 23.67 | 0.000*** |
| Time (in years) | -0.14 | -0.90 | 0.61 | 120.11 | -0.38 | 0.708 |
| Cerebellum | 2.48 | -0.34 | 5.29 | 118.51 | 1.74 | 0.085 |
| Internal Capsule | 0.59 | -2.23 | 3.40 | 118.51 | 0.41 | 0.681 |
| Lesions | -10.46 | -11.43 | -9.49 | 121.77 | -21.36 | 0.000*** |
| NABT | -5.36 | -7.12 | -3.62 | 124.30 | -6.06 | 0.000*** |
| NAGM | -7.89 | -10.61 | -5.19 | 125.74 | -5.77 | 0.000*** |
| Pons | 2.52 | -0.30 | 5.33 | 118.51 | 1.77 | 0.080 |
| Thalamus | -7.79 | -22.44 | 6.84 | 14.79 | -1.12 | 0.282 |
| Whole Brain | -6.37 | -8.66 | -4.10 | 122.33 | -5.53 | 0.000*** |
| Time*Cerebellum | -1.30 | -17.76 | 15.17 | 116.45 | -0.16 | 0.877 |
| Time*Internal Capsule | 1.22 | -15.24 | 17.69 | 116.45 | 0.15 | 0.884 |
| Time*Lesions | 0.59 | -0.30 | 1.49 | 123.10 | 1.32 | 0.191 |
| Time*NABT | 0.14 | -0.85 | 1.14 | 117.80 | 0.28 | 0.778 |
| Time*NAGM | 0.06 | -1.20 | 1.34 | 117.76 | 0.10 | 0.921 |
| Time*Pons | -1.42 | -17.88 | 15.05 | 116.45 | -0.17 | 0.865 |
| Time*Thalamus | -0.21 | -2.50 | 2.09 | 116.85 | -0.18 | 0.859 |
| Time*Whole Brain | 0.14 | -1.28 | 1.55 | 117.51 | 0.19 | 0.850 |
| Number of observations | 145 | | | | | |
| Number of Studies/Subgroups | 27 | | | | | |
| Number of Studies | 13 | | | | | |
| AIC | BIC | loglik | deviance | df. resid | | |
| 676 | 738 | -317 | 634 | 124 | | |

*Supplementary Table 6: Linear mixed-model regression results for longitudinal change in mean MTR in normal-appearing brain tissue.*

| **Linear Mixed Model Fit by Maximum Likelihood (*lmer* package in RStudio)** | | | | | | |
|---|---|---|---|---|---|---|
| **Random effects:** | | | | | | |
| | | | | **Variance** | **SD** | |
| Study/Subgroup | (intercept) | | | 0.076 | 0.28 | |
| Study | (intercept) | | | 38.71 | 6.22 | |
| Residual | | | | 0.040 | 0.20 | |
| **Fixed effects:** | | | | | | |
| ***Predictors*** | ***Estimates*** | ***CI*** | | ***df*** | ***t-value*** | ***p*** |
| (intercept) | 37.55 | 30.85 | 44.25 | 5.01 | 13.48 | 0.000*** |
| Time (in years) | -0.12 | -0.21 | -0.02 | 14.70 | -2.65 | 0.019 |
| Number of observations | 23 | | | | | |
| Number of Studies/Subgroups | 8 | | | | | |
| Number of Studies | 5 | | | | | |
| AIC | BIC | loglik | deviance | df. resid | | |
| 47.8 | 53.5 | -18.9 | 37.8 | 18 | | |

*Supplementary Table 7: Results for a linear mixed regression model assessing change in normal-appearing white matter over time.*

| **Linear Mixed Model Fit by Maximum Likelihood (*lmer* package in RStudio)** | | | | | | |
|---|---|---|---|---|---|---|
| **Random effects:** | | | | | | |
| | | | | **Variance** | **SD** | |
| Study/Subgroup | (intercept) | | | 0.64 | 0.80 | |
| Study | (intercept) | | | 64.79 | 8.05 | |
| Residual | | | | 0.15 | 0.39 | |
| **Fixed effects:** | | | | | | |
| ***Predictors*** | ***Estimates*** | ***CI*** | | ***df*** | ***t-value*** | ***p*** |
| (intercept) | 45.81 | 38.89 | 52.72 | 6.99 | 14.99 | 0.000*** |
| Time (in years) | 0.08 | -0.13 | 0.29 | 31.97 | 0.78 | 0.441 |
| Number of observations | 42 | | | | | |
| Number of Studies/Subgroups | 10 | | | | | |
| Number of Studies | 7 | | | | | |
| AIC | BIC | loglik | deviance | df. resid | | |
| 112.9 | 121.6 | -51.5 | 102.9 | 37 | | |

*Supplementary Table 8: Linear mixed-model results for change in lesion MTR over time.*

| | | | | | | |
|---|---|---|---|---|---|---|
| **Linear Mixed Model Fit by Maximum Likelihood (*lmer* package in RStudio)** | | | | | | |
| **Random effects:** | | | | | | |
| | | | | **Variance** | **SD** | |
| Study/Subgroup | (intercept) | | | 9.27 | 3.04 | |
| Study | (intercept) | | | 39.49 | 6.28 | |
| Residual | | | | 6.17 | 2.48 | |
| **Fixed effects:** | | | | | | |
| ***Predictors*** | ***Estimates*** | ***CI*** | | ***df*** | ***t-value*** | ***p*** |
| (intercept) | 35.21 | 30.54 | 40.05 | 10.10 | 16.04 | 0.000 |
| Time (in years) | 0.38 | -0.56 | 1.30 | 31.08 | 0.82 | 0.420 |
| Number of observations | 49 | | | | | |
| Number of Studies/Subgroups | 19 | | | | | |
| Number of Studies | 10 | | | | | |
| AIC | BIC | loglik | deviance | df. resid | | |
| 286 | 295 | -138 | 276 | 44 | | |

*Supplementary Figure 2: Longitudinal evolution of MTR across different lesion types. APLA: anti-phospholipid antibody status; CEL: contrast-enhancing lesion*

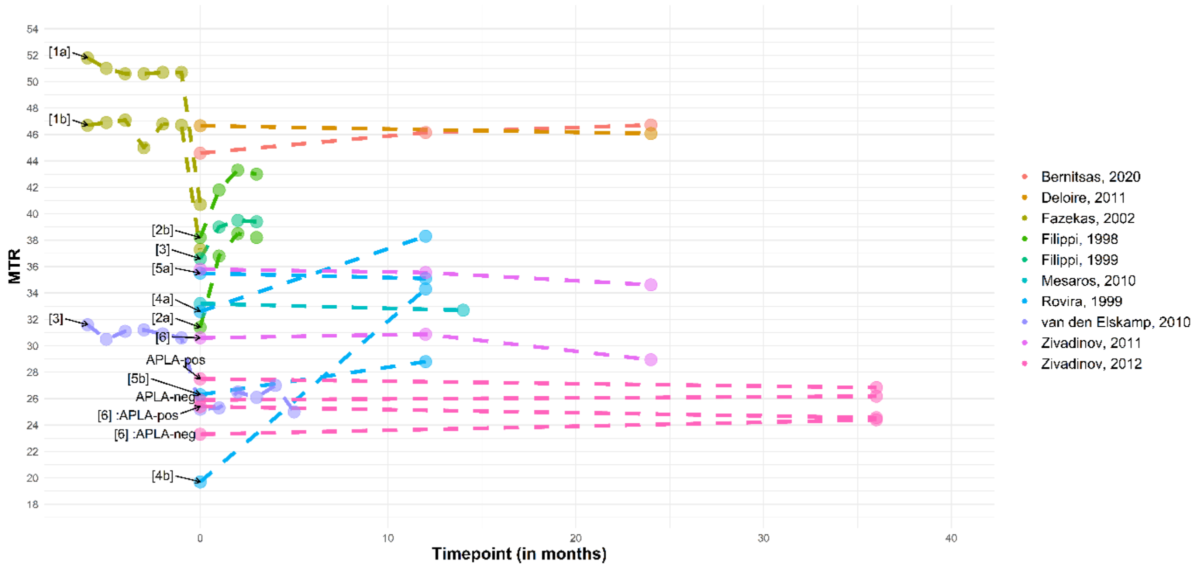

CELs: Contrast-enhancing lesions [1a]: CELs (new, active); [1b]: CELs (reactive); [2a]: CELs (standard dose of Gd);
[2b] CELs (triple dose of Gd) [3]: CELs; [4a]: nodular-enhancing CELs; [4b]: ring-enhancing CELs; [5a]: isointense;
[5b]: hypointense; [6] T1 'black holes'. APLA-pos/neg: anti-phospholipid-positive/negative.
Negative months are shown where study classed contrast-enhancement as timepoint zero.
